# Early childhood linear growth faltering in low- and middle-income countries

**DOI:** 10.1101/2020.06.09.20127001

**Authors:** Jade Benjamin-Chung, Andrew Mertens, John M Colford, Alan E Hubbard, Mark J van der Laan, Jeremy Coyle, Oleg Sofrygin, Wilson Cai, Anna Nguyen, Nolan N Pokpongkiat, Stephanie Djajadi, Anmol Seth, Wendy Jilek, Esther Jung, Esther O Chung, Sonali Rosete, Nima Hejazi, Ivana Malenica, Haodong Li, Ryan Hafen, Vishak Subramoney, Jonas Häggström, Thea Norman, Kenneth H. Brown, Parul Christian, Benjamin F. Arnold, members of the ki Child Growth Consortium

## Abstract

Globally 149 million children under five are estimated to be stunted (length more than 2 standard deviations below international growth standards). Stunting, a form of linear growth faltering, increases risk of illness, impaired cognitive development, and mortality. Global stunting estimates rely on cross-sectional surveys, which cannot provide direct information about the timing of onset or persistence of growth faltering— a key consideration for defining critical windows to deliver preventive interventions. We performed the largest pooled analysis of longitudinal studies in low- and middle-income countries to date (n=32 cohorts, 52,640 children, ages 0-24 months), allowing us to identify the typical age of linear growth faltering onset and to investigate recurrent faltering in early life. The highest incidence of stunting onset occurred from birth to age 3 months. From 0 to 15 months, less than 5% of children per month reversed their stunting status, and among those who did, stunting relapse was common. Early timing and low reversal rates emphasize the importance of preventive intervention delivery within the prenatal and early postnatal phases coupled with continued delivery of postnatal interventions through the first 1000 days of life.

## Introduction

In 2018, 149 million children under 5 years (22% globally) were stunted (length-for-age Z-score >2 standard deviations below the median of the growth standard for age and sex), with the largest burden in South Asia and Africa.^1,2^ Early-life stunting is associated with increased risk of mortality,^3^ diarrhea, pneumonia, and measles in childhood^4,5^ and impaired cognition and productivity in adulthood.^6–8^ Global income would increase by an estimated $176.8 billion per year if linear growth faltering could be eliminated.^9^ The WHO 2025 Global Nutrition Targets^10^ and Sustainable Development Goal 2.2.1 propose to reduce stunting prevalence among children under 5 years from 2012 levels by 40% by 2025.^11^

In low-resource settings, the first 1000 days of life – including the prenatal period – is considered the critical window in which to intervene to prevent stunting.^12^ Intrauterine growth restriction and preterm birth are strongly associated with stunting at 24 months of age.^13^ Most linear growth faltering occurs by age 2 years, and 70% of absolute length deficits by age 5 years occur before age 2 years.^6^ Children who experience linear growth faltering prior to prior to age 2 years can experience catch-up growth at older ages, particularly with improvements to their nutrition, health, and environment.^14–18^ However, the extent of catch-up growth is associated with the timing and extent of early life linear growth faltering.^19^

Granular information about the age of linear growth faltering onset and its persistence in early life will best inform when and how to intervene with preventive measures. Yet, most studies of the global epidemiology of stunting have used nationally representative, cross-sectional surveys – predominantly Demographic Health Surveys (DHS) – to estimate age-specific stunting prevalence.^15,20–22^ Analyses of cross-sectional studies cannot identify longitudinal patterns of linear growth faltering or reversal. Further, they may be subject to survivor bias and fail to include those children most vulnerable to undernutrition. Few studies have estimated age-specific incidence within the first two years of life.^23–27^

We estimated linear growth faltering incidence and reversal and linear growth velocity in 32 longitudinal cohorts in LMICs with multiple, frequent measurements. The analysis provides new insights into the timing of onset and duration of linear growth faltering, with important implications for interventions. We found that linear growth faltering occurs very early in the prenatal and postnatal phase – before age 6 months, when most postnatal linear growth interventions begin. Our findings confirm the importance of the first 1,000 days as a critical window to intervene to prevent linear growth faltering but motivate a renewed focus on prenatal and early postnatal interventions.

### Pooled longitudinal analyses

Here, we report a pooled analysis of 32 longitudinal cohorts from 14 LMICs in South Asia, Sub-Saharan Africa, and Latin America followed between 1987 and 2017. Our objective was to estimate age-specific incidence and reversal of stunting and linear growth velocity from 0 to 24 months. Companion articles report results for child wasting (weight-for-length Z-score < 2 standard deviations below the reference median)^28^ and household, maternal, and child-level risk factors associated with linear growth faltering.^29^ These data were aggregated by the Bill & Melinda Gates Foundation Knowledge Integration (*ki*) initiative and comprise approximately 100 longitudinal studies on child birth, growth and development.^30^ We included cohorts from the database that met five inclusion criteria: 1) conducted in LMICs; 2) had a median year of birth in 1990 or later; 3) enrolled children between birth and age 24 months and measured their length and weight repeatedly over time; 4) did not restrict enrollment to acutely ill children; and 5) collected anthropometry measurements at least every 3 months (Extended Data Fig 1). These criteria ensured we could rigorously evaluate the timing and onset of stunting among children who were broadly representative of general populations in LMICs. Thirty-two cohorts met inclusion criteria, including 52,640 children and 412,458 total measurements from 1987 to 2017 (Fig 1, Extended Data Tables 1-2). Cohorts were located in South Asia (N=17 cohorts in 4 countries), Africa (N=7 in 6 countries), Latin America (N=7 in 3 countries), and Eastern Europe (N=1) (Extended Data Fig 2). 21 cohorts measured children at least monthly, and 11 measured children every 3 months. Cohort sample sizes varied from 119 to 14,074 children. In most cohorts, over 80% of enrolled children had LAZ measurements at each age of measurement (Extended Data Figs 3-4).

**Figure 1.**
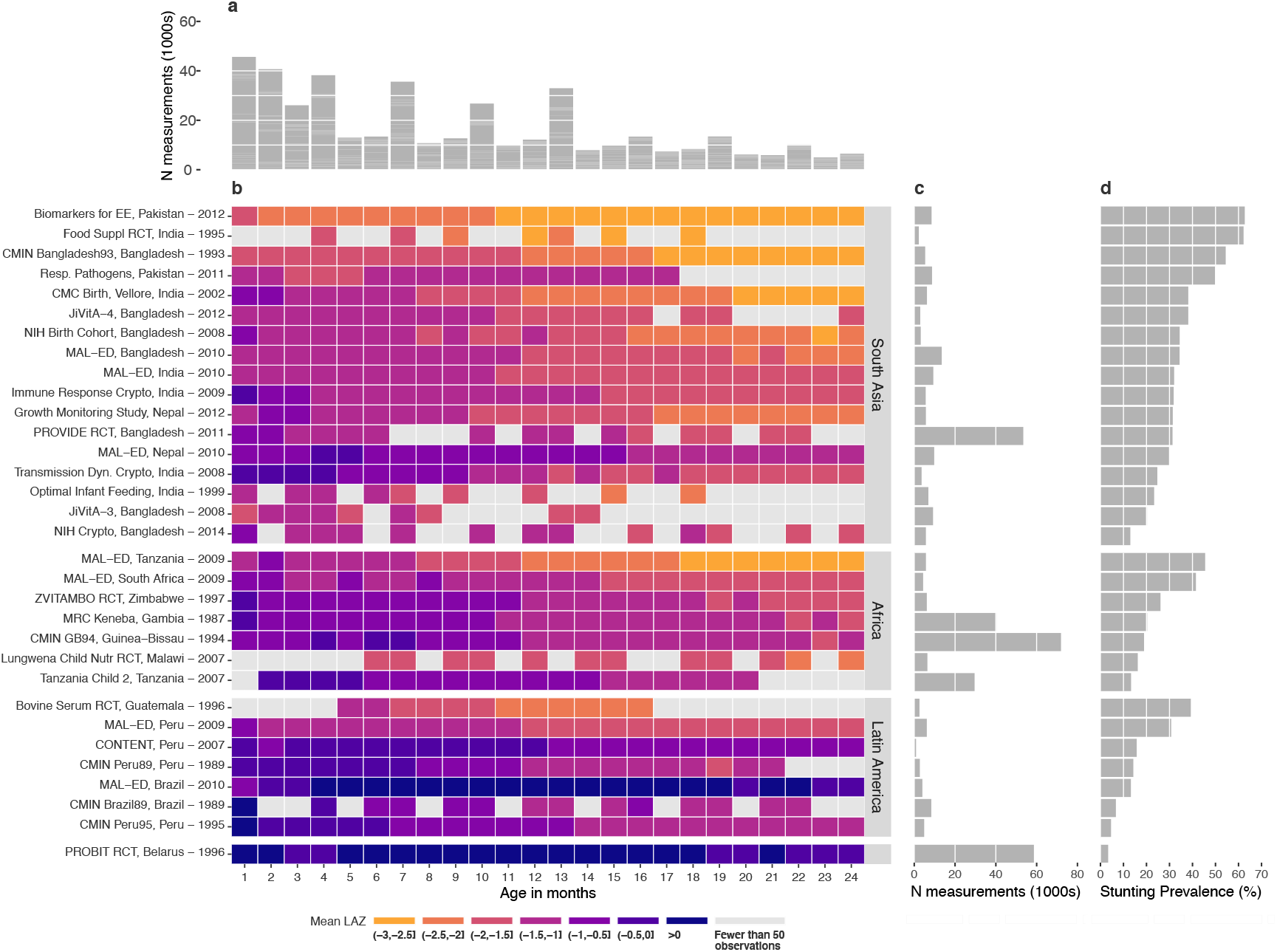
Summaries of included *ki* cohorts. (**a**) Number of observations (1000s) by age in months. (**b**) Mean length-for-age Z-scores by age in months for each cohort. Cohorts are sorted by geographic region and mean length-for-age Z-score. (**c**) Number of observations contributed by each cohort. (**d**) Overall stunting prevalence in each cohort, defined as proportion of measurements with length-for-age z-score < –2.

We calculated length-for-age Z-scores (LAZ) using WHO 2006 growth standards.^31^ We dropped 859 of 413,317measurements (0.2%) because LAZ was unrealistic (> 6 or < –6 Z), and we defined stunting as LAZ < –2 and severe stunting as LAZ < –3.^31^ Unless otherwise indicated, estimates that pool across cohorts used random effects models fit with restricted maximum likelihood estimation.^32,33^ Within each cohort the monthly mean LAZ ranged from –3.06 to +1.31, and the monthly proportion stunted ranged from 0% to 91% (Fig 1).

To assess *ki* cohort representativeness, we compared LAZ from cohorts with LAZ for children 0-24 months of age in contemporary population-based, cross-sectional DHS data in the same countries. *ki* cohorts and DHS Z-score distributions were similar (Fig 2a). Mean LAZ by age was generally lower in *ki* cohorts than in DHS surveys, especially in South Asia, but was slightly higher at certain ages in two Peruvian cohorts (Fig 2b).

**Figure 2.**
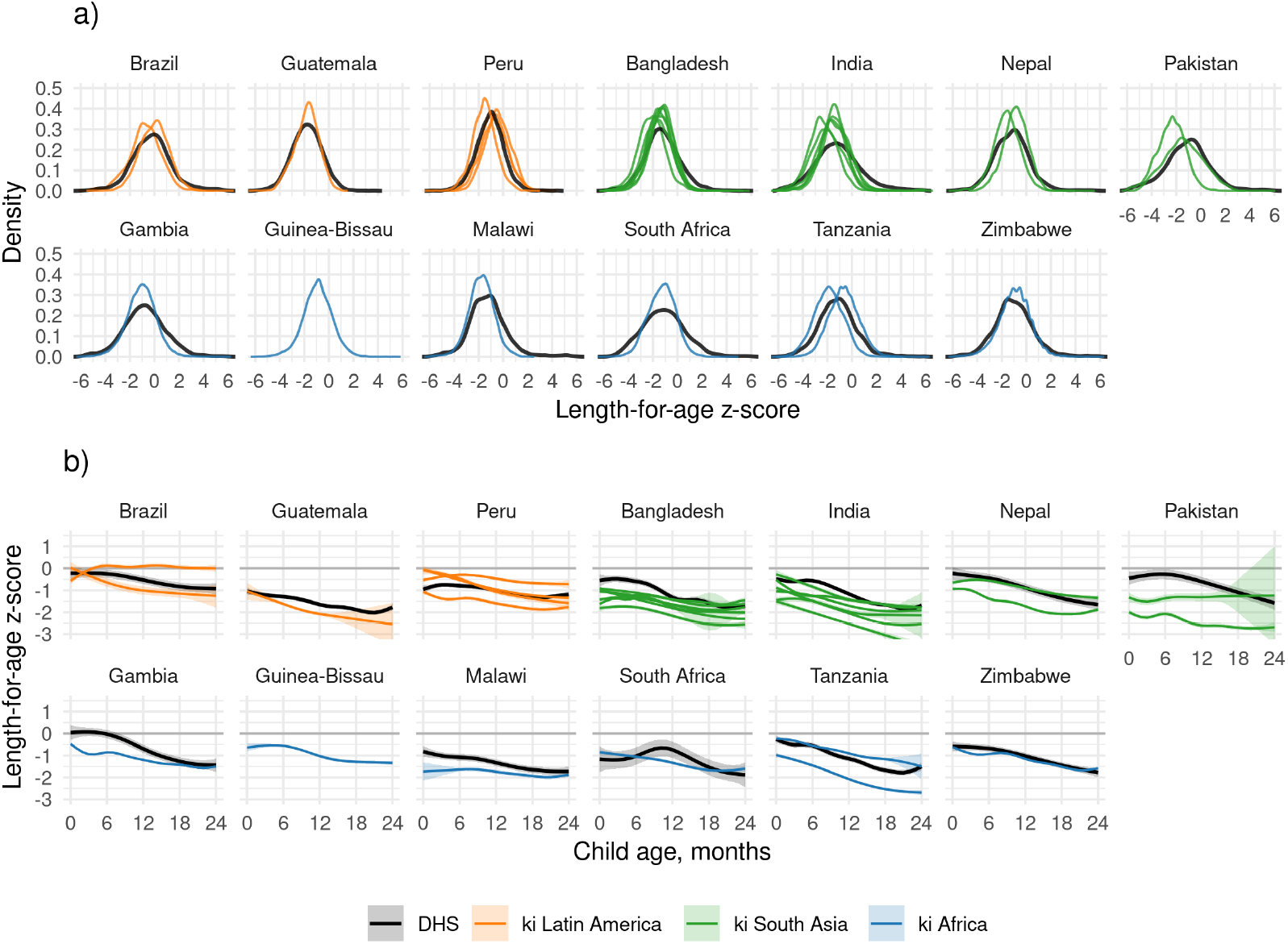
Length-for-age Z-scores by age and region. (**a**) Kernel density distributions of LAZ in DHS in countries that overlap with *ki* cohorts (black lines) and in each *ki* cohort (colored lines). (**b**) Mean length-for-age z-scores (LAZ) by age for Demographic and Health Surveys (DHS) countries overlapping with *ki* cohorts (black lines) and pooled across *ki* longitudinal cohorts with at least quarterly measurement (colored lines) estimated with cubic splines. Shaded bands are approximate 95% simultaneous confidence intervals. The DHS survey was not conducted in Guinea-Bissau during the study period.

### Linear growth faltering as a whole population condition

In approximately half of cohorts, the 95^th^ percentile of the LAZ distribution dropped below 0 by age 15 months (Extended Data Fig 5). This pattern is consistent with the characterization of linear growth faltering as a “whole population” condition.^21^ In most cohorts, as children aged, LAZ distributions shifted downwards (Extended Data Fig 6), and standard deviations and skewness were similar across ages (Extended Data Fig 7).

### Onset of stunting in early life

To measure the timing of stunting onset, we classified a child as a new incident case in three-month age periods if their LAZ dropped below –2 for the first time in that age period. The percentage of children that were stunted at birth ranged from 0.3% to 42% in each cohort and was 13% overall (Fig 3a). The percentage that experienced incident stunting onset between birth and 3 months ranged from 6% to 47% in each cohort and was 16% overall. Children stunted between birth and 3 months accounted for 23% of all children who experienced stunting by age 24 months (69% of children). Very early life stunting onset was most common in South Asia. Trends were similar for severe stunting (https://child-growth.github.io/stunting/severe-stunting.html).

**Figure 3.**
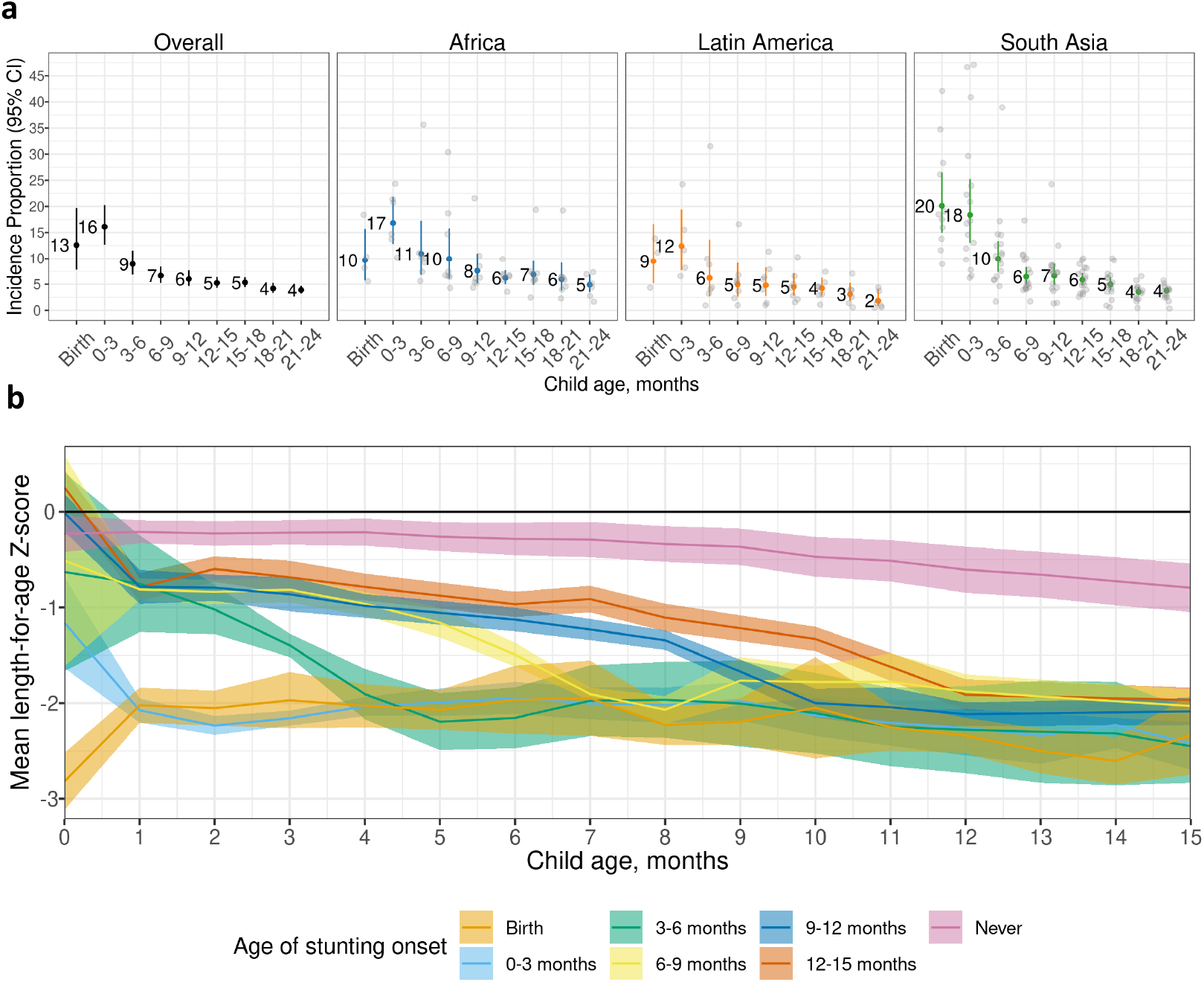
Incidence of stunting and mean LAZ by age. (**a**) Proportion of children experiencing incident stunting onset overall (N=19-32 studies; N= 11,929-42,902 children) and stratified by region (Africa: N=4-8 studies, N = 5,529-15,837 children; Latin America: N=3-7 studies, N=413-1,528 children; South Asia 11-17studies, N=4,514-17,802 children). “0-3” includes age 2 days up to 3 months. Analyses include cohorts with at least quarterly measurements; vertical bars indicate 95% confidence intervals. Gray points indicate cohort-specific estimates. (**b**) Mean length-for-age Z-score (LAZ) stratified by age of incident stunting from birth to age 15 months (N=21 cohorts that measured children at least monthly between birth and age 15 months, N=11,243 children). “Never” includes children who did not become stunted by age 15 months. Shaded ribbons indicate 95% confidence intervals. Pooled results were derived from random effects models with restricted maximum likelihood estimation.

In an exploratory analysis, we stratified age-specific mean LAZ by age of stunting onset among children in monthly measured cohorts and observed three subgroups whose mean LAZ followed statistically different trajectories (Fig 3b). 21% of all children were born with mean LAZ < –1, and their mean LAZ stabilized around –2 from age 1 month onward, with differences at birth in this group narrowing over time, likely via regression to the mean. 14% of children were born with mean LAZ between –1 and 0 at birth, and in these children, mean LAZ approached –2 at subsequent ages. The remaining 65% never met the criteria for stunting; yet, among these children, mean LAZ was between – 0.5 and 0 at birth, and it declined steadily, reaching close to –1 by age 15 months. A companion article reports characteristics that increase risk of earlier versus later growth faltering.^30^

### Stunting reversal and relapse

We hypothesized that 1) lower than average linear growth (LAZ <0) would persist among children who experienced stunting reversal (i.e., LAZ increased from below –2 to above –2), and 2) children who experienced stunting reversal would experience stunting relapse at later ages. To test these hypotheses, we classified a child’s change in stunting status from birth to 15 months among monthly-measured cohorts (measurement frequency beyond 15 months was less consistent). The proportion previously stunted who were either reversed or no longer stunted at 15 months was 16% (Fig 4a). New incidence of stunting was highest at birth and declined steadily to 3.3% per month by age 4 months (Fig 4b). Incidence rates of new and relapse stunting exceeded rates of reversal at all ages (Fig 4b). The proportion with stunting relapse following reversal ranged from 2-3.5% from ages 6 to 15 months. Within each cohort, 0.5% to 10% of children experienced stunting reversal each month, and 0.1% to 11% of experienced stunting relapse each month (Fig 4b).

**Figure 4.**
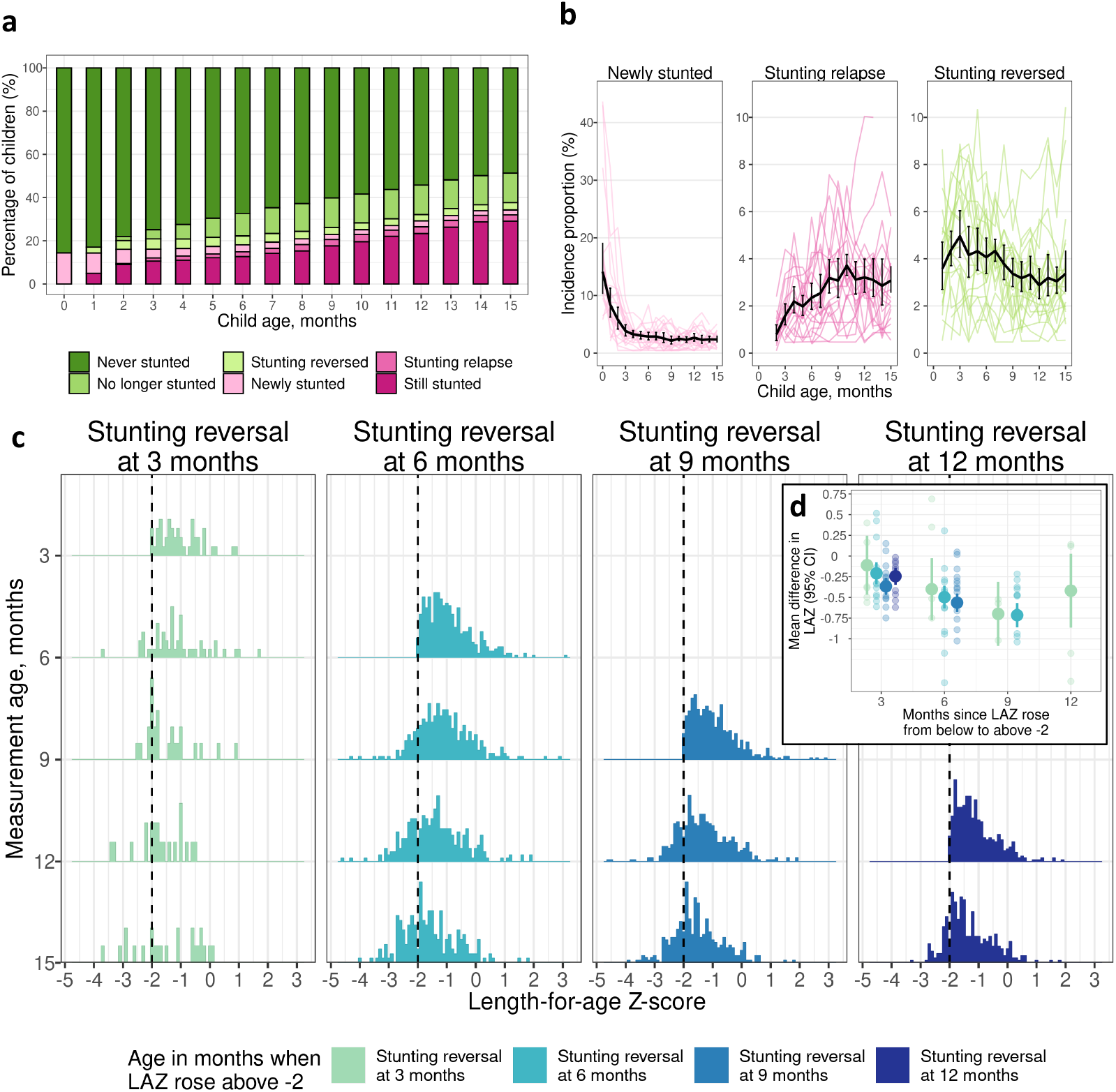
Stunting reversal and relapse. (**a**) Percentage of children with stunting reversal and relapse by age. (**b**) Incidence proportion of new stunting, stunting relapse, and stunting reversal by age. The black line presents estimates pooled using random effects with restricted maximum likelihood estimation. Colored lines indicate cohort-specific estimates. (**c**) Distribution of LAZ at subsequent measurements among children who experienced stunting reversal. (**d**) Mean difference in LAZ following stunting reversal at each subsequent age of measurement compared to the age of reversal. Panels a, c, d include data from 21 cohorts in 10 countries with at least monthly measurement (N =11,424). All panels contain data up to age 15 months because in most cohorts, measurements were less frequent above 15 months.

Among children who experienced stunting reversal, we summarized the LAZ distribution at older ages. We then estimated the mean difference in LAZ measured at older ages compared to when stunting was reversed. At the time of stunting reversal, the LAZ distribution mode was close to the –2 cutoff (Fig 4c, Extended Data Fig 8). As children aged, LAZ distributions gradually shifted downwards, illustrating that linear growth deficits continued to accumulate. Among children who experienced stunting reversal before 6 months, mean difference in LAZ 9 months later was -0.69 (95% CI -0.84, -0.55; cohort-specific range: -1.04, -0.22) (Fig 4d). Children who were older at the time of reversal experienced larger decline in subsequent LAZ compared to younger children (Fig 4d). Overall, improvements in LAZ following stunting reversal were neither sustained nor large enough to erase linear growth deficits and did not resemble a biological recovery process for most children.

### Growth velocity by age and sex

We defined linear growth velocity as the change in length between two time points divided by the number of months between the time points (cm/month). We also estimated within-child rates of LAZ change per month, which compares changes in a child’s length relative to the WHO standard over time. After 6 months, changes in length and LAZ within-child were minimal. From 0-3 months, cohort-specific length velocity ranged from below the 1^st^ percentile of the WHO standard to above the 50^th^ for boys and above the 75^th^ percentile for girls (Fig 5a). At subsequent ages, length velocity in each cohort was mostly between the 15^th^ and 50^th^ percentiles of the WHO standard, except in one cohort in Belarus, which had higher length velocity. Larger deficits at the youngest ages were consistent with highest incidence of stunting from birth to age 3 months (Fig 3a). The difference in LAZ within child per month was largest from 0-3 months; after age 3 months, the mean change in LAZ within child was <0.3 between different age intervals (Fig 5b). Generally, velocity within age was higher in Latin America than in South Asia and Africa (Extended Data Fig 8).

**Figure 5.**
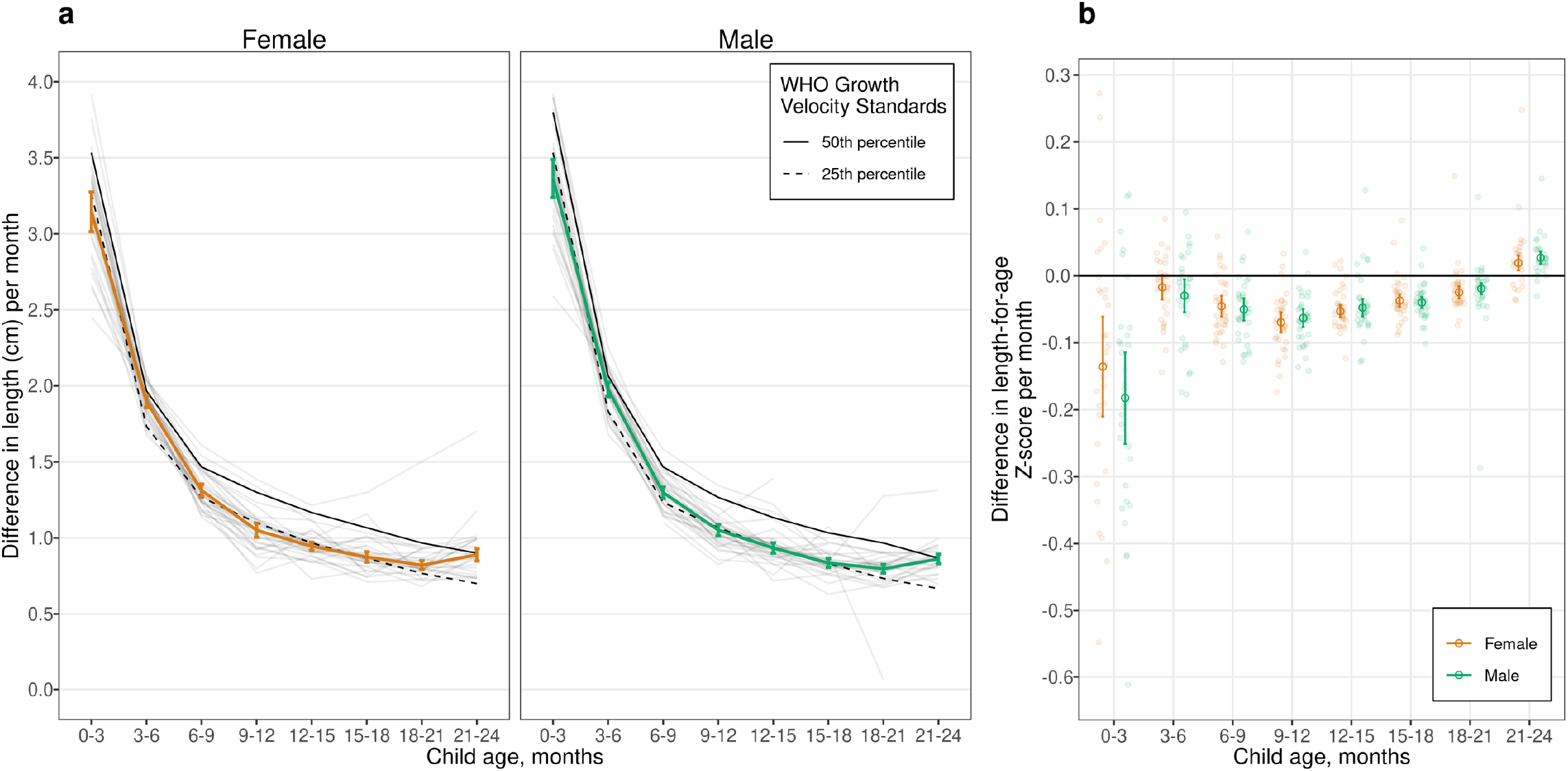
Linear growth velocity by age and sex. (**a**) Within-child difference in length in centimeters per month stratified by age among male (green line) and female (orange line) children; 25^th^ percentile of the WHO Growth Velocity Standards (dashed black lines); and the 50^th^ percentile (solid black line). Light gray lines indicate cohort-specific linear growth velocity curves. (**b**) Within-child difference in length-for-age Z-score per month by age and sex. Smaller partially transparent points indicate cohort-specific estimates. Both panels include 32 *ki* cohorts in 14 countries that measured children at least quarterly (n = 52,640 children) pooled using random effects models fit with restricted maximum likelihood estimation. Vertical bars indicate 95% confidence intervals.

## Discussion

This large-scale analysis of 32 longitudinal cohorts from LMICs revealed new insights into the timing, persistence, and recurrence of linear growth faltering from birth to age 2 years. Prior cross-sectional studies found that stunting prevalence increased gradually with age.^15,20–22^ In contrast, we found that overall, 13% of children were born stunted, 25% of children experienced stunting onset between birth and 6 months, and 31% experienced onset from 6 to 24 months (Fig 3, Fig 4b). Most children who experienced stunting reversal continued to experience linear growth deficits, and over 20% were stunted again at later measurements (Fig 4). Even among children who were never met criteria for stunting before age 2 years, mean LAZ steadily declined with age (Fig 3b). Our findings reinforce that linear growth faltering among children in LMICs is a whole-population phenomenon, with both stunted and not stunted children experiencing suboptimal growth trajectories in early life.^21^

Our findings that 13% of children were stunted at birth and that the birth prevalence was 20% in South Asia underscores the importance of pre-pregnancy and prenatal interventions to reduce stunting, especially in South Asia where at-birth stunting was 24%. These interventions include maternal micronutrient and macronutrient supplementation,^34,35^ increasing women’s autonomy and education,^36^ reducing adolescent pregnancies in LMICs by delaying the age of marriage and first pregnancy,^37^ and promoting family planning.^38^ Interventions to prevent prenatal infections, such as intermittent preventive treatment for malaria, may also increase fetal linear growth in regions where such infections co-occur with linear growth faltering.^39^

In this study, 25% of children became stunted between birth and age 6 months, yet few child nutrition interventions are recommended by the World Health Organization in this age range. In the neonatal period, those interventions include delayed cord clamping, neonatal vitamin K administration, and kangaroo mother care.^40^ Beyond the neonatal period, the sole recommended intervention is exclusive breastfeeding,^40^ which significantly reduces the risk of mortality and morbidity but has not been found to reduce infant stunting.^4,41–44^ Additional research is needed to identify interventions that prevent linear growth faltering between birth and 6 months, including nutritional support of the lactating parent and the vulnerable infant.^45^ Interventions may need to focus on upstream risk factors, such as maternal pre-conception and prenatal health and nutrition, and microbiota.

We found that 31% of children became stunted during complementary feeding (age 6-24 months). Meta-analyses evaluating the effectiveness of interventions during this phase on linear growth have reported modest impacts of lipid-based nutrient supplements,^46^ modest or no impact of micronutrient supplementation,^47^ and no impact of water and sanitation improvements, deworming, or maternal education.^47^ The dearth of effective postnatal interventions to improve linear growth motivates renewed efforts to identify alternative, possibly multisectoral interventions, and to improve intervention targeting and implementation.^48,49^

There were several limitations to the analyses. First, length estimates may be subject to measurement error; stunting reversal and relapse analyses that rely on thresholds are more sensitive to such errors. However, detailed assessments of measurement quality indicated that measurement quality was high across cohorts (https://child-growth.github.io/stunting/QA.html). Second, estimates of LAZ at birth using the WHO Child Growth Standards overestimate stunting in preterm infants. Accurate estimates of gestational age were not available in included cohorts; seven cohorts measured gestational age by recall of last menstrual period or newborn examination, and one cohort measured gestational age by ultrasound. In a sensitivity analysis adjusting for gestational age pooling across cohorts that measured it, stunting prevalence at birth was 1% lower (Extended Data Fig 9). Third, included cohorts were not inclusive of all countries in the regions presented here, and linear growth faltering was more common in included African and South Asian cohorts than in corresponding contemporary representative surveys. The consistency between attained linear growth patterns in this and nationally representative DHS surveys (Fig 2) suggests that overall, our results have reasonably good external validity. For growth velocity, the cohorts represented populations close to the 25th percentile of international standards (Figure 5a). Fourth, the included cohorts measured child length every 1-3 months, and ages of measurement varied, so different numbers of children and cohorts contributed to each estimate. However, when we repeated analyses in cohorts with monthly measurements from birth to 24 months (n=18 cohorts in 10 countries, 10,830 children), results were similar (https://child-growth.github.io/stunting/monthly.html). Finally, our inferences are limited to the first two years of life since very few included studies measured children at older ages. Other studies, however, have found that stunting status in early life is associated with health outcomes later in life, and the timing and extent of early life linear growth faltering is associated with the magnitude of later catch up growth.^6–8,16,17,19^

### Conclusion

Current WHO 2025 Global Nutrition Targets and Sustainable Development Goal 2.2.1 aim to reduce stunting prevalence among children under 5 years by 2025. Our findings suggest that defining stunting targets at earlier ages (e.g., stunting by 3 or 6 months) would help focus attention on the period when interventions may be most impactful. In addition, our results motivate renewed efforts to identify effective prenatal and early postnatal interventions and improve delivery of early life interventions to prevent linear growth faltering among children in LMICs.

## Data Availability

The data that support the findings of this study are available from the Bill and Melinda Gates Foundation Knowledge Integration project upon reasonable request.

## Materials and Methods

### Study designs and inclusion criteria

We included all longitudinal observational studies and randomized trials available through the *ki* project on April 2018 that met 5 inclusion criteria (Extended Data Fig 1).

1. *Studies that were conducted in low- or middle-income countries*. Children in these countries have the largest burden linear growth faltering and are the key target population for preventive interventions.
2. *Studies that had a median year of birth in 1990 or later*. This restriction resulted in a set of studies spanning the period from 1987 to 2017 and excluded older studies that are less applicable to current policy dialogues.
3. *Studies that enrolled children between birth and age 24 months and measured their length and weight repeatedly over time*. We were principally interested in growth faltering during the first 1,000 days (including gestation), thought to be the key window for linear growth faltering.
4. *Studies that did not restrict enrollment to acutely ill children*. Our focus on descriptive analyses led us to target, to the extent possible, the general population. We thus excluded some studies that exclusively enrolled acutely ill children, such as children who presented to hospital with acute diarrhea or who were severely malnourished.
5. *Studies that collected anthropometry measurements at least every 3 months* to ensure that we adequately captured incident episodes and recovery.

Thirty-two longitudinal cohorts in 14 countries followed between 1987 and 2017 met inclusion criteria. There was no evidence of secular trends in LAZ (https://child-growth.github.io/stunting/secular-trends.html). We calculated cohort measurement frequency as the median days between measurements. If randomized trials found effects on growth within the intervention arms, the analyses were limited to the control arm. We included all measurements under 24 months of age, assuming months were 30.4167 days. We excluded extreme measurements of LAZ > 6 or < –6 following WHO growth standard recommendations.^1^ In many studies, investigators measured length shortly after birth because deliveries were at home, but the majority of measurements were within the first 7 days of life (https://child-growth.github.io/stunting/age-meas.html); for this reason, we grouped measurements in the first 7 days as birth measurements. Gestational age was measured in only eight cohorts (7 cohorts measured it by recall of last menstrual period or newborn examination; 1 measured it by ultrasound); thus, we did not attempt to exclude preterm infants from the analyses.

### Quality assurance

The *ki* data team assessed the quality of individual cohort datasets by checking the range of each variable for outliers and values that are not consistent with expectation. Z-scores were calculated using the median of replicate measurements and the 2006 WHO Child Growth Standards.^1^ In a small number of cases a child had two anthropometry records at the same age, in which case we used the mean of the records. Analysts reviewed bivariate scatter plots to check for expected correlations (e.g., length by height, length/height/weight by age, length/height/weight by corresponding Z-score). Once the individual cohort data was mapped to a single harmonized dataset, analysts conducted an internal peer review of published articles for completeness and accuracy. Analysts contacted contributing investigators to seek clarification about potentially erroneous values in the data and revised the data as needed.

### Outcome definitions

We used the following summary measures in the analysis:

<u>Incident stunting episodes</u> were defined as a change in LAZ from above –2 Z in the prior measurement to below –2 Z in the current measurement. Similarly, we defined severe stunting episodes using the –3 Z cutoff. Children were considered at risk of stunting at birth, so children born stunted were considered to have an incident episode of stunting at birth. Children were also assumed to be at risk of stunting at the first measurement in non-birth cohorts and trials. Children whose first measurement occurred after birth were assumed to have experienced stunting onset at the age halfway between birth and the first measurement. The vast majority of children were less than 5 days of age at their first measurement life (https://child-growth.github.io/stunting/age-meas.html).

<u>Incidence proportion</u> We calculated the incidence proportion of stunting during a defined age range (e.g. 3-6 months) as the proportion of children at risk of becoming stunted who became stunted during the age range (the onset of new episodes).

<u>Changes in stunting status</u> were classified using the following categories: “Never stunted”: children with LAZ ≥–2 at previous ages and the current age. “No longer stunted”: children who previously reversed their stunting status with LAZ ≥–2 at the current age. “Stunting reversal”: children with LAZ <–2 at the previous age and LAZ ≥ –2 at the current age. “Newly stunted”: children whose LAZ was previously always ≥–2 and with LAZ <–2 at the current age. “Stunting relapse”: children who were previously stunted with LAZ ≥–2 at the previous age and LAZ <–2 at the current age. “Still stunted”: children whose LAZ was <–2 at the previous and current age.

<u>Growth velocity</u> was calculated as the change in length in centimeters between two time points divided by the number of months between the time points. We compared the change in length in centimeters per month measures to the WHO Child Growth Standards for linear growth velocity.^2^ We also estimated within-child rates of change in LAZ per month.

### Measurement frequency

Analyses of incidence and growth velocity (Figs 3 and 5) included cohorts with at least quarterly measurements in order to include as many cohorts as possible. Analyses of stunting reversal (Fig 4) were restricted to cohorts with at least monthly measurements to allow evaluation of changes in stunting status with higher resolution.

### Subgroups of interest

We stratified the above outcomes within the following subgroups: child age, grouped into one- or three-month intervals, (depending on the analysis); the region of the world (Asia, sub-Saharan Africa, Latin America); child sex, and the combinations of those categories.

### Statistical analysis

All analyses were conducted in R version 3.4.2.^3^

#### Estimation of mean LAZ by age in Demographic and Health Surveys and *ki* cohorts

We downloaded standard DHS individual recode files for each country from the DHS program website (https://dhsprogram.com/). We used the most recent standard DHS datasets for the individual women’s, household, and height and weight datasets from each country. We obtained variables for country code, sample weight, cluster number, primary sampling unit, and design stratification from the women’s individual survey recode files. From the height and weight dataset, we used standard recode variables corresponding to the 2006 WHO growth standards for height-for-age.

After excluding missing observations, restricting to measurements of children 0-24 months of age, and restricting to z-scores within WHO-defined plausible values, surveys collected from 1996 to 2018 in countries that overlapped with *ki* cohorts with the exception of Guinea-Bissau because the DHS survey was not conducted there during the study period (Extended Data Table 3).

We classified countries into regions (South Asia, Latin America, and Africa) using the World Health Organization regional designations with the exception of the classification for Pakistan, which we included in South Asia to be consistent with prior linear growth studies using DHS.^4^ One included cohort was from Belarus, and we chose to exclude it from region-stratified analyses as it was the only European study.

We estimated age-stratified mean from ages 0 to 24 months within each DHS survey, accounting for the complex survey design and sampling weights. We then pooled estimates of mean LAZ for each age in months across countries using a fixed effects estimator (details below). We computed two sets of pooled results: 1) DHS measuring children 0-24 months in countries that overlapped with *ki* study countries and 2) all DHS countries measuring children 0-24 months in each geographic region (as in Fig 2) and (https://child-growth.github.io/stunting/DHS.html). We compared DHS estimates with mean LAZ by age in the ki study cohorts, which we estimated using penalized cubic-splines with bandwidth chosen using generalized cross-validation.^5^ We used splines to estimate age-dependent mean LAZ in the *ki* study cohorts to smooth any age-dependent variation in the mean caused by less frequently measured cohorts.

#### Distribution models

To investigate how the mean, standard deviation, and skewness of LAZ distributions varied by age, we fit linear models with skew-elliptical error terms using maximum likelihood estimation. We fit models separately by cohort.

#### Fixed and random effects models

Several analyses pooled results across study cohorts. We estimated each age-specific mean using a separate estimation and pooling step. We first estimated the mean in each cohort, and then pooled age-specific means across cohorts, while allowing for a cohort-level random effect. This approach enabled us to include the most information possible for each age-specific mean, while accommodating slightly different measurement schedules across the cohorts. Each cohort’s data only contributed to LAZ or stunting incidence estimates at the ages for which it contributed data.

The primary method of pooling was using random effects models. This modeling approach assumes that studies are randomly drawn from a hypothetical population of longitudinal studies that could have been conducted on children’s linear growth in the past or future. We also fit fixed effects models as a sensitivity analysis (https://child-growth.github.io/stunting/fixed-effects.html); inferences about estimates from fixed effects models are restricted to only the included studies.^6^

Random effects models assume that the true population outcomes *θ* are normally distributed with mean *μ* and variance *τ*^2^ (i.e., that *θ* ∼ N(*μ, τ*^2^)). To estimate outcomes in this study, the random effects model is defined as follows for each study in the set of *i* = 1, …, *k* studies:

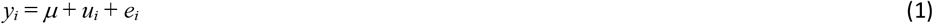

where *y*_*i*_ is the observed outcome in study *i, u*_*i*_ is the random effect for study *i*, and *e*_*i*_ is the estimated outcome for study *i*, and *e*_*i*_ is the sampling error within study *i*. The model assumes that *u*_*i*_ ∼ N(0, *τ*^2^) and *e*_*i*_ ∼ N(0, *v*_*i*_), where *v*_*i*_ is the study-specific sampling variance. We fit random effects models using the restricted maximum likelihood estimator.^7,8^ If a model failed to converge, models were fit using a maximum likelihood estimator instead. The estimate of *μ* is the estimated mean outcome in the hypothetical population of studies (i.e., the estimated outcome pooling across study cohorts).

We also fit inverse variance weighted fixed effects models defined as follows:

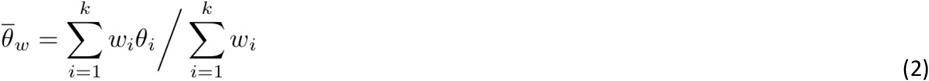

where 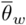 is the weighted mean outcome in the set of *k* included studies, and *w*_*i*_ is a study-specific weight, defined as the inverse of the study-specific sampling variance *v*_*i*_. 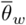 is the estimated mean outcome in the specific studies included in this analysis.

For both types of outcomes, we pooled binary outcomes on the logit scale and then back-transformed estimates after pooling to constrain confidence intervals between 0 and 1. While the probit transformation more closely resembles common distributions for physiologic variables, in practice the logit transformation produces nearly identical estimates and is more convenient for estimation. For cohort-stratified analyses, which did not pool across studies, we estimated 95% confidence intervals using the normal approximation (https://child-growth.github.io/stunting/cohort.html).

#### Estimation of incidence

We estimated incidence as defined above in 3-month age intervals within specific cohorts and pooled within region and across all studies (Fig 3). Pooled analyses used random effects models for the primary analysis and fixed effects models for sensitivity analyses as described above.

#### Estimation of changes in stunting status

To assess fluctuations in stunting status over time, we conducted an analysis among cohorts with at least monthly measurements from birth through age 15 months to provide sufficient granularity to capture changes in stunting status. We estimated the proportion of children in each stunting category defined above under “Changes in stunting status” at each month from birth to 15 months. To ensure that percentages summed to 100%, we present results that were not pooled using random effects. Analyses using random effects produced similar results (https://child-growth.github.io/stunting/fixed-effects.html#changes-in-stunting-status-by-age).

To examine the distribution of LAZ among children with stunting reversal, we created subgroups of children who experienced stunting reversal at ages 3, 6, 9, and 12 months and then summarized the distribution of the children’s LAZ at ages 6, 9, 12, and 15 months. Within each age interval, we estimated the mean difference in LAZ at older ages compared to the age of stunting reversal and estimated 95% confidence intervals for the mean difference. Pooled analyses used random effects models for the primary analysis and fixed effects models for sensitivity analyses as described above.

#### Linear growth velocity

We estimated linear growth velocity within 3-month age intervals stratified by sex, pooling across study cohorts (Fig 5) as well as stratified by geographic region (Extended Data Fig 9) and study cohort (https://child-growth.github.io/stunting/cohort.html#length-velocity-by-age-and-sex). Analyses included cohorts that measured children at least quarterly. We included measurements within a two-week window around each age in months to account for variation in the age of each length measurement. Pooled analyses used random effects models for the primary analysis and fixed effects models for sensitivity analyses as described above (https://child-growth.github.io/stunting/fixed-effects.html#linear-growth-velocity-1).

### Sensitivity Analyses

We conducted three sensitivity analyses. First, to assess whether inclusion of PROBIT, the single European cohort, influenced our overall pooled inference, we repeated analyses excluding the PROBIT cohort. Results were very similar with and without the PROBIT cohort (https://child-growth.github.io/stunting/exclude-PROBIT.html). Second to explore the influence of differing numbers of cohorts contributing data at different ages, we conducted a sensitivity analysis in which we subset data to cohorts that measured anthropometry monthly from birth to 24 months (n=21 cohorts in 10 countries, 11,424 children (https://child-growth.github.io/stunting/monthly.html). Third, we compared estimates pooled using random effects models presented in the main text with estimates pooled using fixed effects inverse variance weighted models. The random effects approach was more conservative in the presence of study heterogeneity (https://child-growth.github.io/stunting/fixed-effects.html).

### Data and code availability

The data that support the findings of this study are available from the Bill and Melinda Gates Foundation Knowledge Integration project upon reasonable request. Replication scripts for this analysis are available here: https://github.com/child-growth/ki-longitudinal-growth.

## Acknowledgments

This research was financially supported by a global development grant (OPP1165144) from the Bill & Melinda Gates Foundation to the University of California, Berkeley, CA, USA. We would also like to thank the following collaborators on the included cohorts and trials for their contributions to study planning, data collection, and analysis: Muhammad Sharif, Sajjad Kerio, Ms. Urosa, Ms. Alveen, Shahneel Hussain, Vikas Paudel (Mother and Infant Research Activities), Anthony Costello (University College London), Benjamin Torun, Lindsey M Locks, Christine M McDonald, Roland Kupka, Ronald J Bosch, Rodrick Kisenge, Said Aboud, Molin Wang, Azaduzzaman, Abu Ahmed Shamim, Rezaul Haque, Rolf Klemm, Sucheta Mehra, Maithilee Mitra, Kerry Schulze, Sunita Taneja, Brinda Nayyar, Vandana Suri, Poonam Khokhar, Brinda Nayyar, Poonam Khokhar, Jon E Rohde, Tivendra Kumar, Jose Martines, Maharaj K Bhan, and all other members of the study staffs and field teams. We would also like to thank all study participants and their families for their important contributions.

## Author contributions

Conceptualization: J.B., A.M., J.M.C., K.H.B., P.C., B.F.A

Funding Acquisition: J.M.C., A.E.H., M.J.V., B.F.A.

Data curation: J.B., A.M., J.C., O.S., W.C., A.N., N.N.P., W.J., E.J., E.O.C., S.D., N.H., I.M., H.L., R.H., V.S., J.H., T.N.

Formal analyses: J.B., A.M., J.C., O.S., W.C., A.N., N.N.P., W.J., E.J., E.O.C., S.D., N.H., I.M., H.L., V.S., B.F.A

Methodology: J.B., A.M., J.M.C, J.C., O.S., N.H., I.M., A.E.H., M.J.V.,K.H.B., P.C., B.F.A.

Visualization: J.B., A.M., A.N., N.N.P., S.D., A.S., J.C., R.H., E.J., K.H.B., P.C., B.F.A.

Writing – Original Draft Preparation: J.B., A.M., B.F.A.

Writing – Review & Editing: J.B., A.M., J.M.C., K.H.B., P.C., B.F.A., *ki* Child Growth Consortium members

## Additional information

Supplementary Information is available for this paper here: https://child-growth.github.io/stunting/

Correspondence and requests for materials should be addressed to Jade Benjamin-Chung and Benjamin F. Arnold.

## Manuscript figures

**Extended Data Figure 1.**
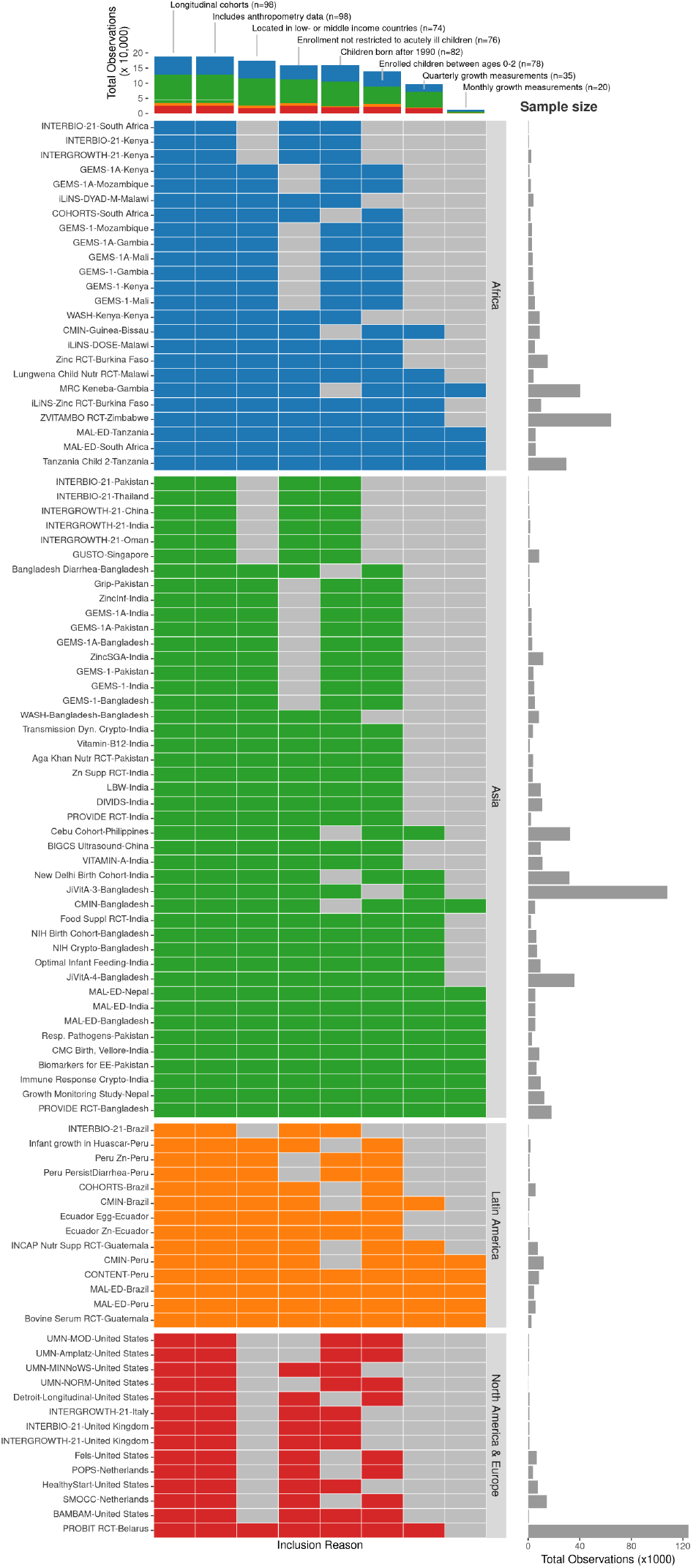
*ki* cohort selection. Analyses focused on longitudinal cohorts to enable the estimation of prospective incidence rates and growth velocity. In April 2018, there were 86 longitudinal studies on GHAP. From this set, we applied five inclusion criteria to select cohorts for analysis. Our rationale for each criterion follows. (1) Studies were conducted in lower income or middle-income countries. (2) Studies had a median year of birth in 1990 or later. (3) Studies measured length and weight between birth and age 24 months. (4) Studies did not restrict enrollment to acutely ill children. (5) Studies collected anthropometry measurements at least every 3 months. Each colored cell indicates a criterion that was met. For studies that met all inclusion criteria, all cells in their row are colored. The bars at the top of the plot show the number of observations in each study that met each inclusion criterion by region.

**Extended Data Figure 2.**
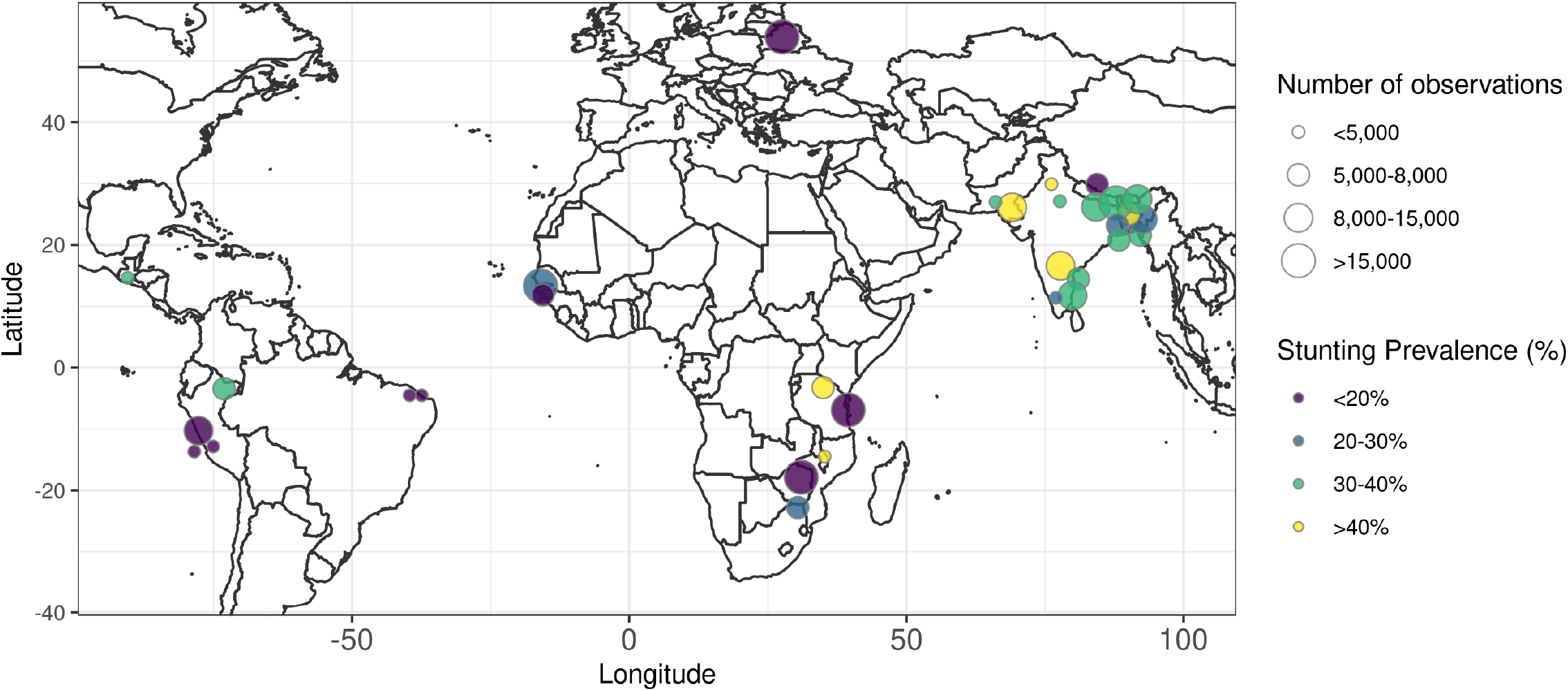
Stunting prevalence by geographic location of *ki* cohorts. Locations are approximate, represented as nation-level centroids and jittered slightly for display. The size of each centroid indicates the number of observations contributing to each estimate. The color of each centroid indicates the level of stunting prevalence.

**Extended Data Figure 3.**
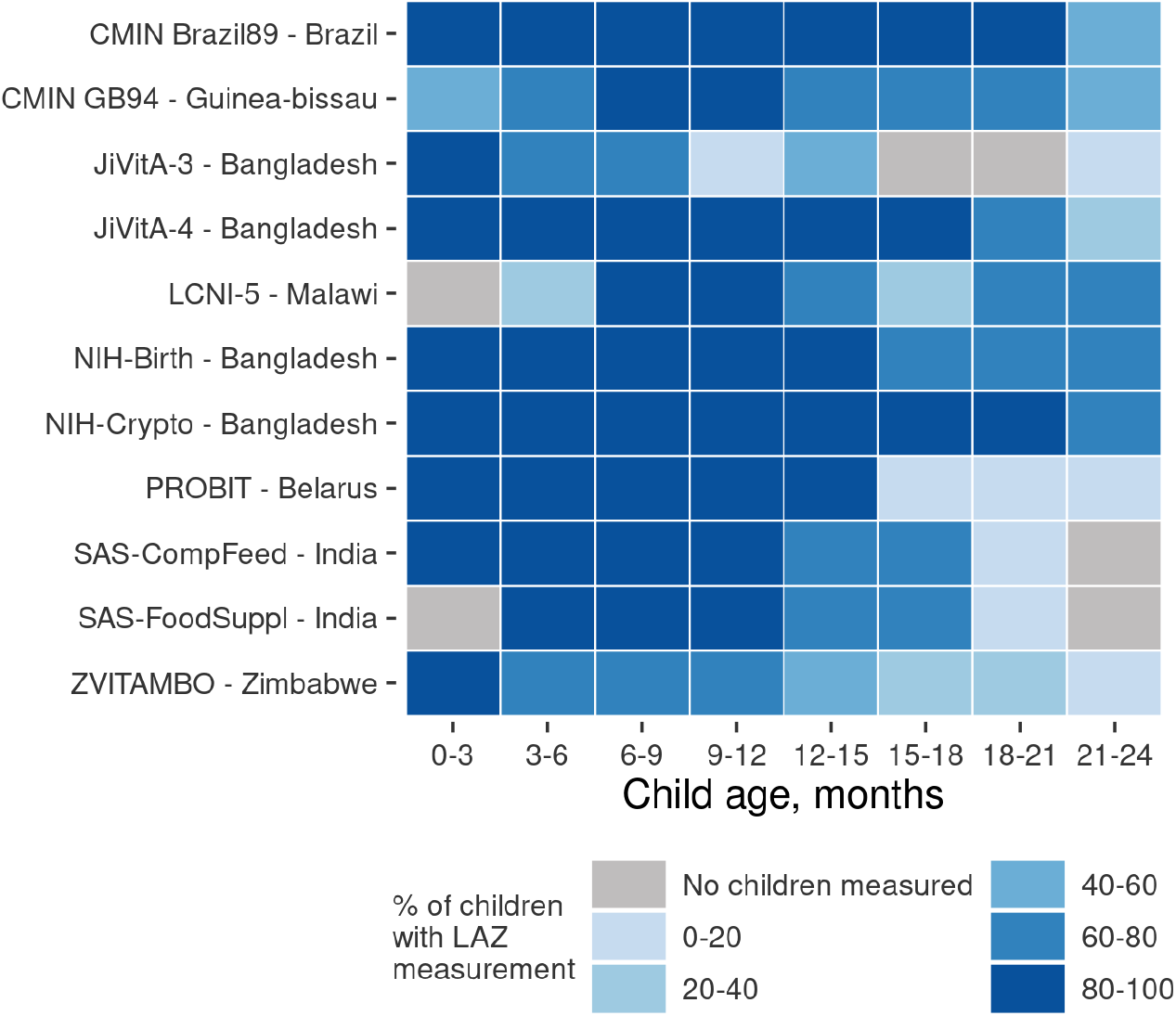
Percentage of enrolled children measured in each *ki* cohort with quarterly measurements.

**Extended Data Figure 4.**
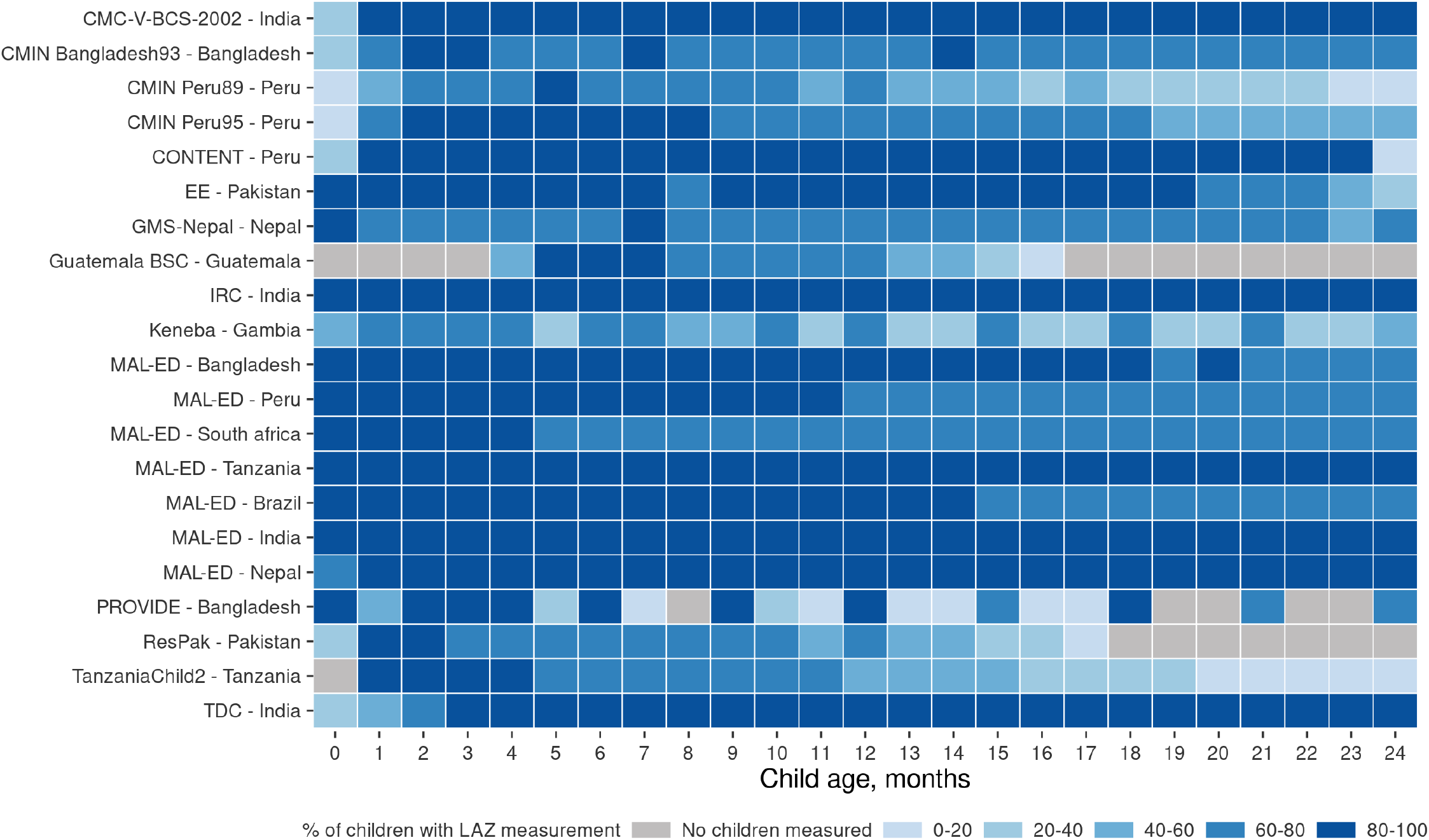
Percentage of enrolled children measured in each *ki* cohort with monthly measurements.

**Extended Data Figure 5.**
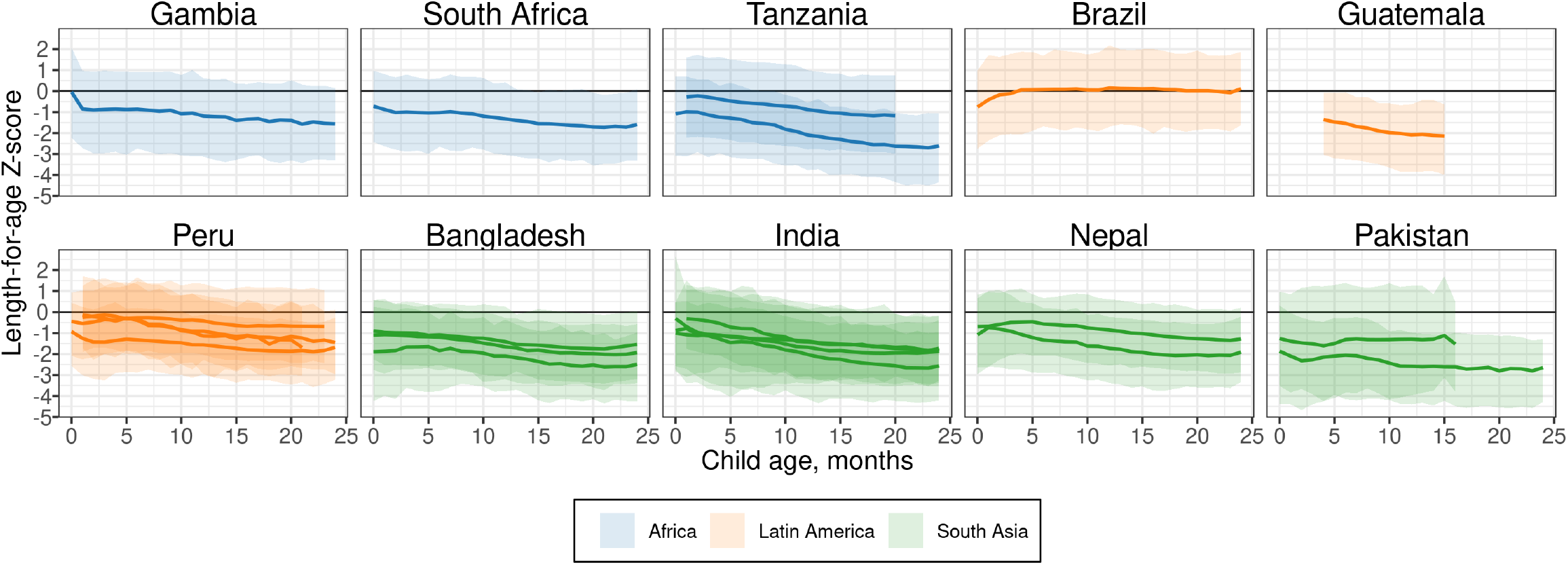
Mean, 5th and 95^th^ percentile of length-for-age Z-score by age in *ki* longitudinal cohorts estimated with cubic splines in cohorts with at least monthly measurement. The shaded bands span the 5^th^ to the 95^th^ percentile of length-for-age Z-score in each cohort. The solid line indicates the mean in each cohort at each age (N=21 cohorts that measured children at least monthly, N=11,424 children).

**Extended Data Figure 6.**
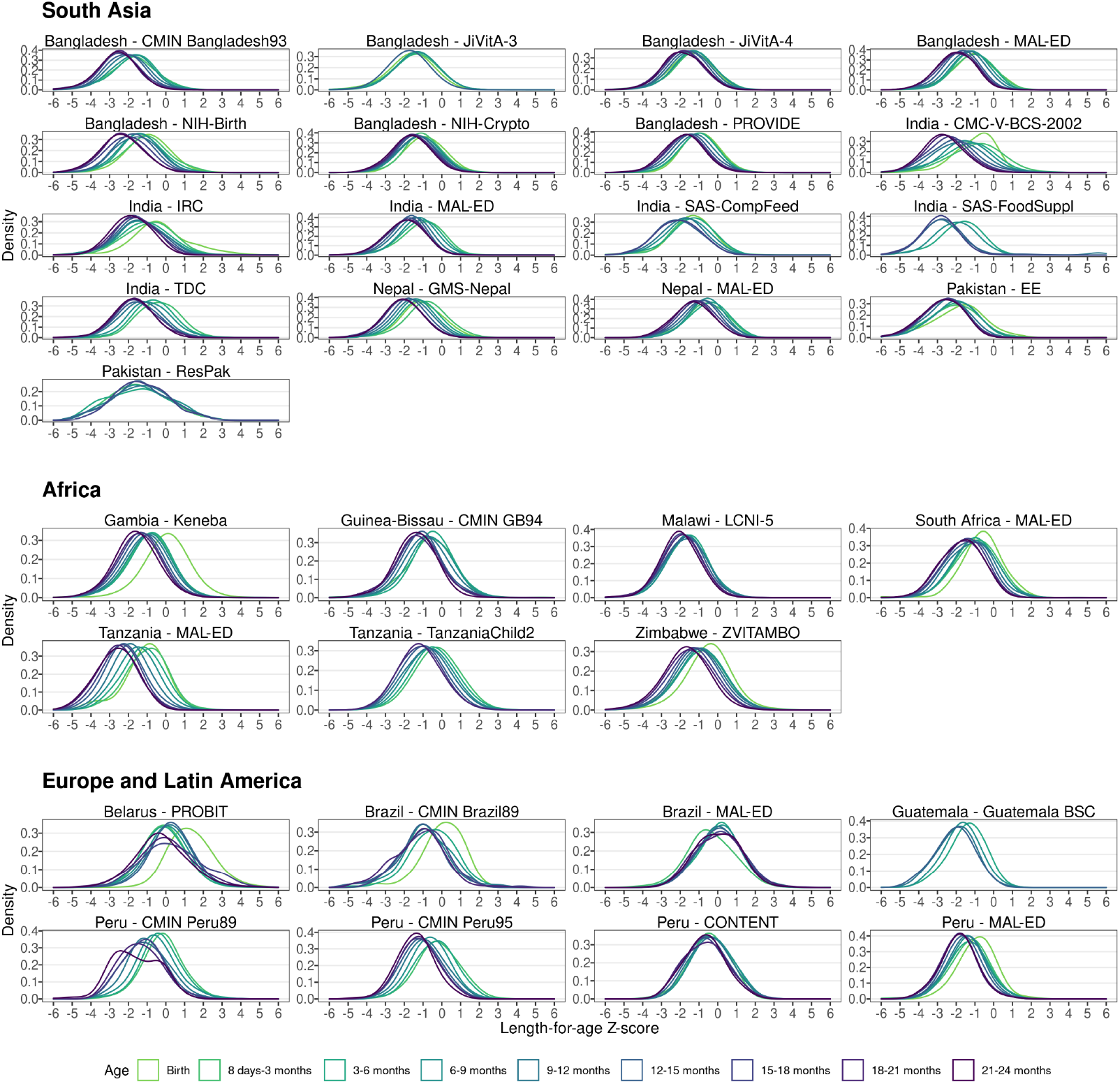
Kernel density of length-for-age Z-score by age and cohort. In South Asia, includes data from 17 cohorts, <u>21,</u>223 children, and <u>159,</u>884 measurements. In Africa, includes 7 cohorts, <u>21,</u>671 children, and <u>164,</u>431 measurements. In Europe and Latin America, includes 8 cohorts, 9,746 children, and 88,143 measurements.

**Extended Data Figure 7.**
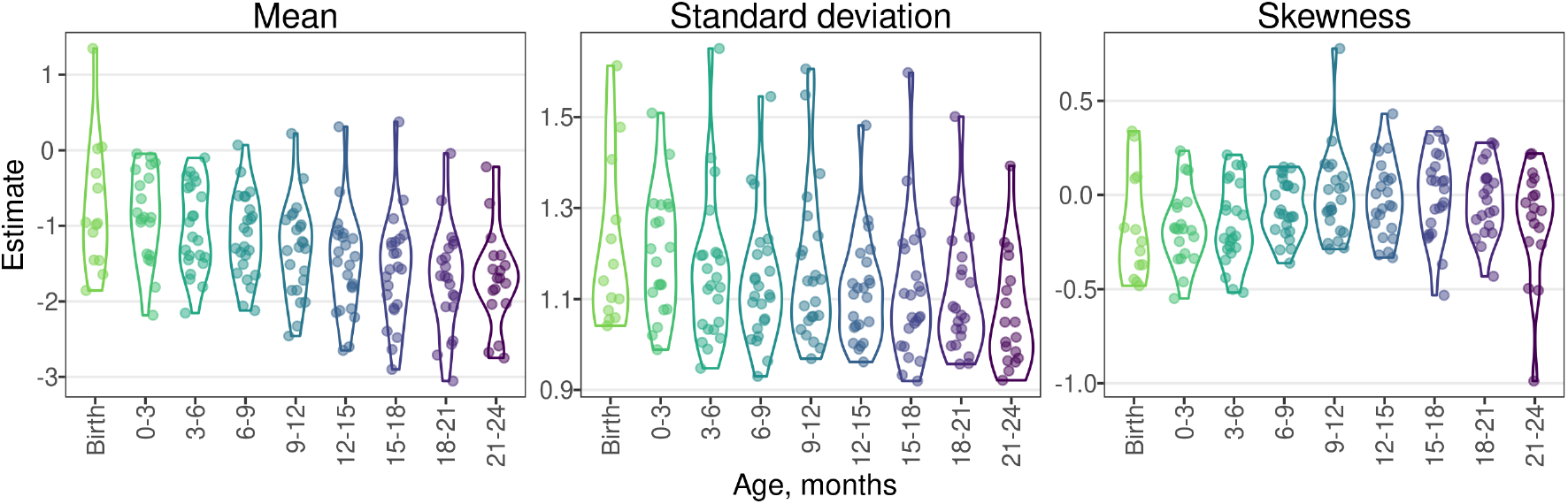
Parametric mean, standard deviation, and Pearson’s index of skewness estimates by age and cohort. Estimates were obtained from linear models with skew-elliptical error terms fit using maximum likelihood estimation. Includes 412,458 measurements from 52,640 children in 32 cohorts.

**Extended Data Figure 8.**
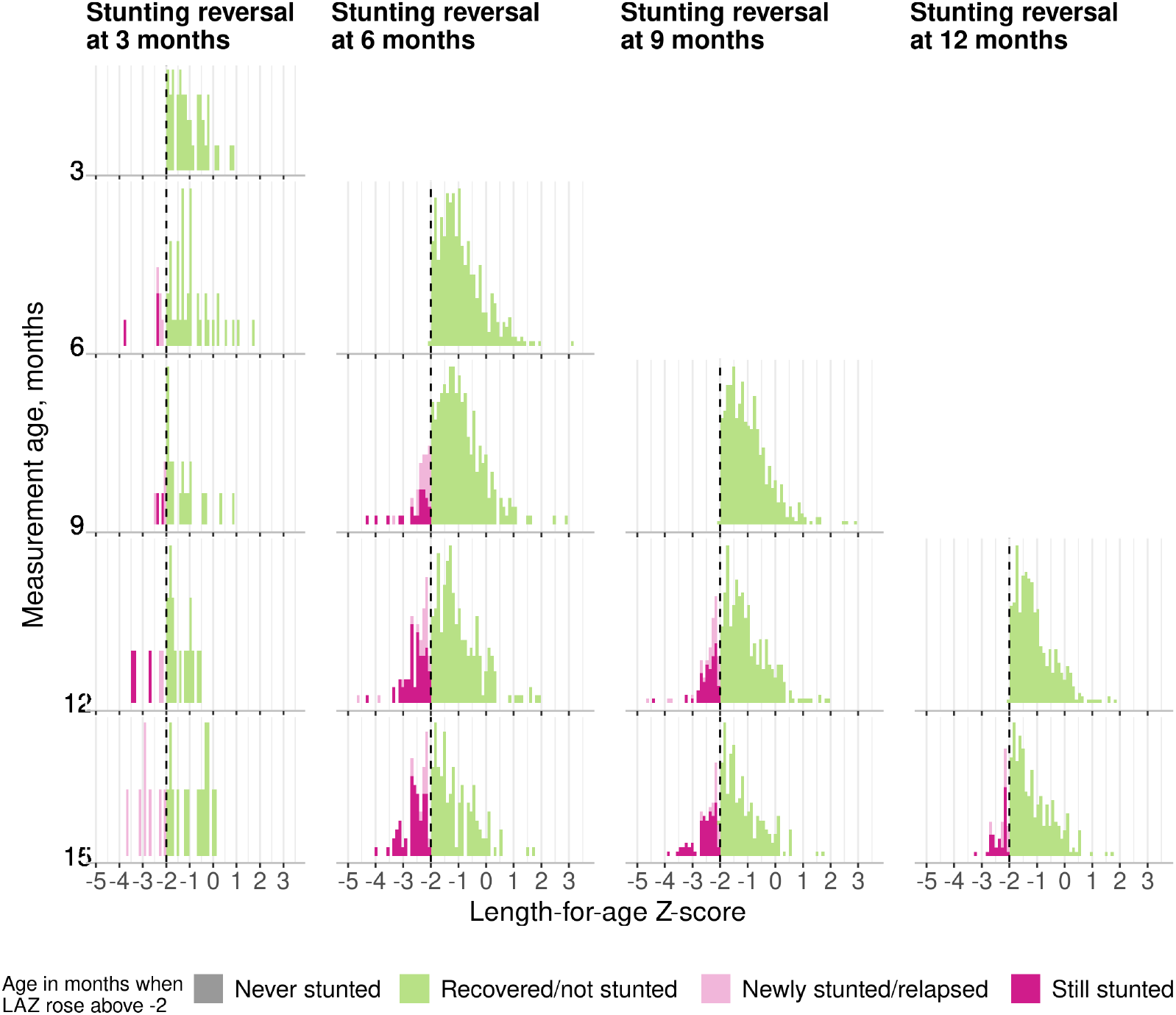
Distribution of LAZ at subsequent measurements after stunting reversal. Includes data from 21 cohorts in 10 countries with at least monthly measurement (N = 11,424 children). All panels contain data up to age 15 months because in most cohorts, measurements were less frequent above 15 months.

**Extended Data Figure 9.**
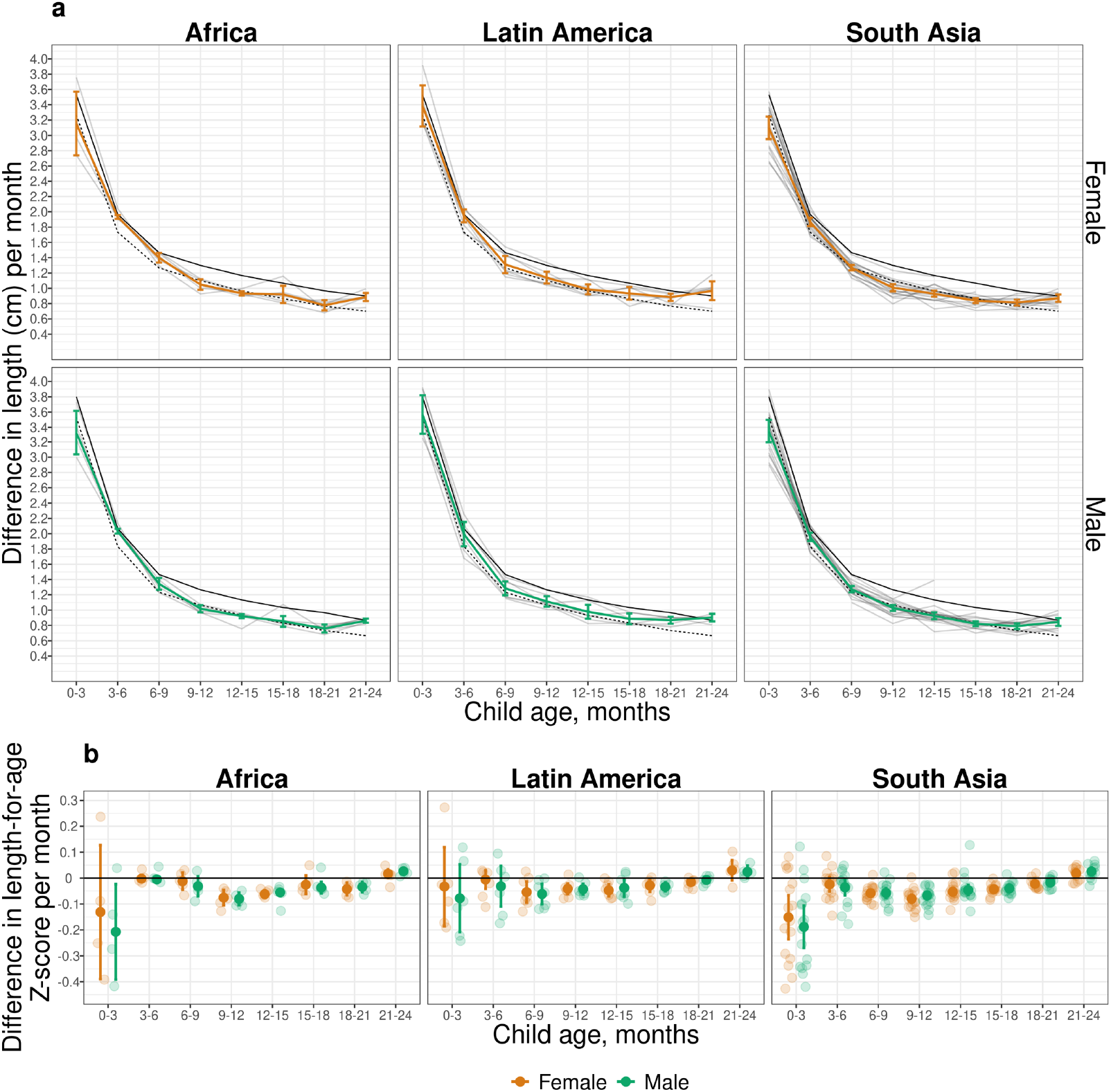
Linear growth velocity by age and sex stratified by region. **a**) Within-child difference in length in centimeters per month stratified by age, sex, and region. Dashed black line indicates 25^th^ percentile of the WHO Growth Velocity Standards; solid black line indicates the 50^th^ percentile. Colored lines indicate and vertical bars indicate 95% confidence intervals for *ki* cohorts. Light gray lines indicate cohort-specific linear growth velocity curves. (**b**) Within-child difference in length-for-age Z-score per month by age, sex, and region. Smaller partially transparent points indicate cohort-specific estimates. Results shown in all panels were derived from 32 *ki* cohorts in 14 countries that measured children at least quarterly (n = 52,640 children).

**Extended Data Figure 10.**
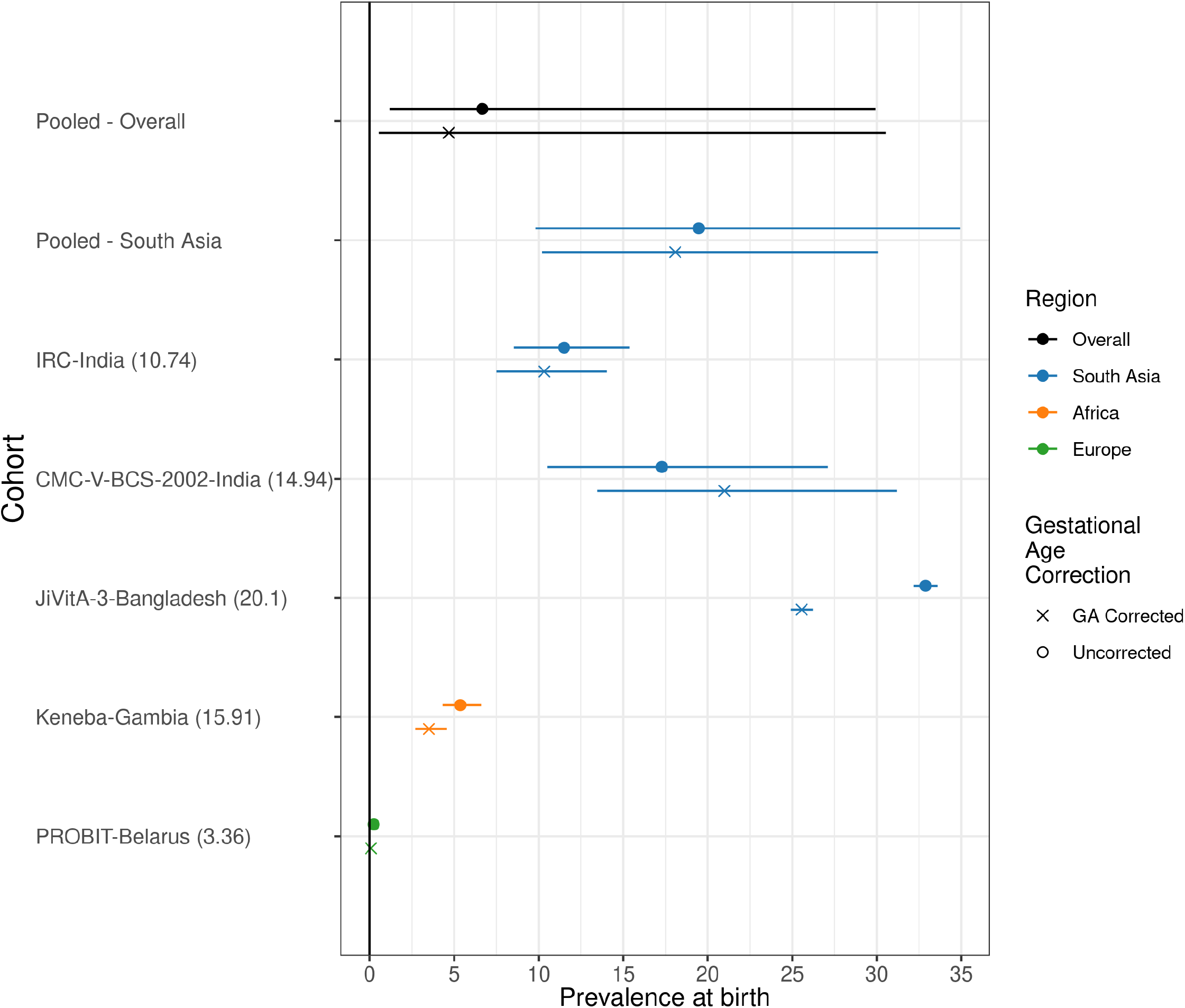
Comparison of stunting prevalence at birth with and without gestational age correction. This figure includes the results from correcting at-birth Z-scores in the *ki* cohorts that measured gestational age (GA) for 37,218 measurements in 5 cohorts. The number in the parentheses following each cohort name indicates the prevalence of pre-term birth in each cohort. The corrections are using the Intergrowth standards and are implemented using the R *growthstandards* package (https://ki-tools.github.io/growthstandards/). Overall, the stunting prevalence at birth decreased slightly after correcting for gestational age, but the cohort-specific results are inconsistent. Observations with GA outside of the Intergrowth standards range (<168 or > 300 days) were dropped for both the corrected and uncorrected data. Prevalence increased after GA correction in some cohorts due to high rates of late-term births based on reported GA. Gestational age was estimated based on mother’s recall of the last menstrual period in the Jivita-3, IRC, and CMC-V-BCS-2002 cohorts, was based on the Dubowitz method (newborn exam) in the Keneba cohort, and was based on ultrasound measurements in the PROBIT trial.

**Extended Data Table 1.** Summary of *ki* cohorts.

| Region, Study ID | Country | Study Years | Design | Children Enrolled* | Anthropometry measurement ages (months) | Total measurements* | Primary References |
| --- | --- | --- | --- | --- | --- | --- | --- |
| **South Asia** |  |  |  |  |  |  |  |
| Biomarkers for EE | Pakistan | 2013-2015 | Prospective cohort | 379 | Birth, 1, 2, ..., 18 | 8484 | Iqbal et al 2018 Nature Scientific Reports<sup>1</sup> |
| Resp. Pathogens | Pakistan | 2011 - 2014 | Prospective cohort | 284 | Birth, 1, 2, ..., 17 | 3177 | Ali et al 2016 Journal of Medical Virology<sup>2</sup> |
| Growth Monitoring Study | Nepal | 2012 - Ongoing | Prospective cohort | 698 | Birth, 1, 2, ..., 24 | 13465 | Not yet published |
| MAL-ED | Nepal | 2010 - 2014 | Prospective cohort | 240 | Birth, 1, 2, ..., 24 | 5703 | Shrestha et al 2014 Clin Infect Dis<sup>3</sup> |
| CMC Birth Cohort, Vellore | India | 2002 - 2006 | Prospective cohort | 373 | Birth, 0.5, 1, 1.5, ..., 24 | 8709 | Gladstone et al. 2011 NEJM<sup>4</sup> |
| MAL-ED | India | 2010 - 2012 | Prospective cohort | 251 | Birth, 1, 2, ..., 24 | 5702 | John et al 2014 Clin Infect Dis<sup>5</sup> |
| Vellore Crypto Study | India | 2008 - 2011 | Prospective cohort | 410 | Birth, 1, 2, ..., 24 | 9771 | Kattula et al. 2014 BMJ Open<sup>6</sup> |
| CMIN 93 | Bangladesh | 1993 - 1996 | Prospective Cohort | 277 | Birth, 3, 6, ..., 24 | 5363 | Pathela et al 2007 Acta Paediatrica<sup>7</sup> |
| TDC | India | 2008-2011 | Quasi-experimental | 160 | Birth, 1, 2, ..., 24 | 3593 | Sarkar et al. 2013 BMC Public Health<sup>8</sup> |
| MAL-ED | Bangladesh | 2010 - 2014 | Prospective cohort | 265 | Birth, 1, 2, ..., 24 | 5604 | Ahmed et al 2014 Clin Infect Dis<sup>9</sup> |
| PROVIDE RCT | Bangladesh | 2011 - 2014 | Individual RCT | 700 | Birth, 6, 10, 12, 14, 17, 18, 24, 39, 40, 52, 53 (weeks) | 9207 | Kirkpatrick et al 2015 Am J Trop Med Hyg<sup>10</sup> |
| Food Suppl RCT | India | 1995 - 1996 | Individual RCT | 418 | Baseline, 6, 9, 12 | 2232 | Bhandari et al 2001 J Nutri<sup>11</sup> |
| Optimal Infant Feeding | India | 1999 - 2001 | Cluster RCT | 472 | Birth, 3, 6, ..., 18 | 2948 | Bhandari et al 2004 J Nutri<sup>12</sup> |
| NIH Birth Cohort | Bangladesh | 2008 - 2009 | Prospective Cohort | 629 | Birth, 3, 6, ..., 12 | 6215 | Korpe et al. 2016 PLOS NTD<sup>13</sup> |
| JiVitA-4 Trial | Bangladesh | 2012 - 2014 | Cluster RCT | 1434 | 6, 9, 12, 14, 18 | 9344 | Christian et al 2015 IJE<sup>14</sup> |
| JiVitA-3 Trial | Bangladesh | 2008 - 2012 | Cluster RCT | 13475 | Birth, 1, 3, 6, 12, 24 | 53453 | West et al JAMA 2014<sup>15</sup> |
| NIH Cryptosporidium Study | Bangladesh | 2014 - 2017 | Prospective cohort | 758 | Birth, 3, 6, ..., 24 | 6914 | Steiner et al 2018 Clin Infect Dis<sup>16</sup> |
| **Africa** |  |  |  |  |  |  |  |
| MAL-ED | Tanzania | 2009 - 2014 | Prospective cohort | 262 | Birth, 1, 2, ..., 24 | 5713 | Mduma et al 2014 Clin Infect Dis<sup>17</sup> |
| Tanzania Child 2 | Tanzania | 2007 - 2011 | Individual RCT | 2396 | 1, 2, ..., 20 | 29565 | Locks et al Am J Clin Nutr 2016<sup>18</sup> |
| MAL-ED | South Africa | 2009 - 2014 | Prospective cohort | 314 | Birth, 1, 2, ..., 24 | 6163 | Bessong et al 2014 Clin Infect Dis<sup>19</sup> |
| MRC Keneba | Gambia | 1987 - 1997 | Cohort | 2915 | Birth, 1, 2, ..., 24 | 40275 | Schoenbuchner et al. 2019, AJCN<sup>20</sup> |
| ZVITAMBO Trial | Zimbabwe | 1997 - 2001 | Individual RCT | 14074 | Birth, 6 wks, 3, 6, 9, 12 | 71928 | Malaba et al 2005 Am J Clin Nutr<sup>21</sup> |
| Lungwena Child Nutrition RCT | Malawi | 2011 - 2014 | Individual RCT | 840 | Birth, 1-6 wk, 6, 12 18 | 4336 | Mangani et al. 2015, Mat Child Nutr<sup>22</sup> |
| iLiNS-Zinc Study | Burkina Faso | 2010 - 2012 | Cluster RCT | 797 | 9, 12, 15, 18 | 1471 | Hess et al 2015 Plos One<sup>23</sup> |
| CMIN GB94 | Guinea Bissau | 1994 - 1997 | Prospective Cohort | 870 | Enrollment and every 3 months after | 6451 | Valentiner-Branth 2001 Am J Clin Nutr<sup>24</sup> |
| **Latin America** |  |  |  |  |  |  |  |
| MAL-ED | Peru | 2009 - 2014 | Prospective cohort | 303 | Birth, 1, 2, ..., 24 | 6142 | Yori et al 2014 Clin Infect Dis<sup>25</sup> |
| CONTENT | Peru | 2007 - 2011 | Prospective cohort | 215 | Birth, 1, 2, ..., 24 | 8339 | Jaganath et al 2014 Helicobacter<sup>26</sup> |
| Bovine Serum RCT | Guatemala | 1997 - 1998 | Individual RCT | 315 | Baseline, 1, 2, ..., 8 | 2545 | Begin et al. 2008, EJCN<sup>27</sup> |
| MAL-ED | Brazil | 2010 - 2014 | Prospective cohort | 233 | Birth, 1, 2, ..., 24 | 4858 | Lima et al 2014 Clin Infect Dis<sup>28</sup> |
| CMIN Brazil89 | Brazil | 1989-2000 | Prospective Cohort | 119 | Birth, 1, 2, ..., 24 | 889 | Moore et al. 2001 Int J Epidemiol.<sup>29</sup> |
| CMIN Peru95 | Peru | 1995 - 1998 | Prospective Cohort | 224 | Birth, 1, 2, ..., 24 | 3979 | Checkley et al. 2003 Am J Epidemiol.<sup>30</sup> |
| CMIN Peru89 | Peru | 1989 - 1991 | Prospective Cohort | 210 | Birth, 1, 2, ..., 24 | 2742 | Checkley et al. 1998 Am J Epidemiol.<sup>31</sup> |
| **Europe** |  |  |  |  |  |  |  |
| PROBIT Study | Belarus | 1996 - 1997 | Cluster RCT | 8127 | 1, 2, 3, 6, 9, 12 | 58649 | Kramer et al 2001 JAMA<sup>32</sup> |
| *Children enrolled is for children with measurements under 2 years of age. Total measurements are number of measurements of anthropometry on children under 2 years of age. |  |  |  |  |  |  |  |

**Extended Data Table 2.**

|  | CMC-V-BCS-2002, India | CMIN<br>Bangladesh<br>93 | CMIN Brazil<br>89 | CMIN Guinea-Bissau 94 | CMIN Peru<br>89 | CMIN Peru<br>95 | CONTENT,<br>Peru | EE, Pakistan | GMS-Nepal | Guatemala<br>BSC |
| --- | --- | --- | --- | --- | --- | --- | --- | --- | --- | --- |
|  | (N=373) | (N=280) | (N=119) | (N=885) | (N=210) | (N=224) | (N=215) | (N=380) | (N=698) | (N=315) |
| Sex |  |  |  |  |  |  |  |  |  |  |
| Female | 187 (50.1%) | 122 (43.6%) | 62 (52.1%) | 443 (50.1%) | 100 (47.6%) | 96 (42.9%) | 109 (50.7%) | 185 (48.7%) | 328 (47.0%) | 162 (51.4%) |
| Male | 186 (49.9%) | 158 (56.4%) | 57 (47.9%) | 442 (49.9%) | 110 (52.4%) | 128 (57.1%) | 106 (49.3%) | 195 (51.3%) | 370 (53.0%) | 153 (48.6%) |
| Birthweight |  |  |  |  |  |  |  |  |  |  |
| Mean (SD) | 2910 (434) | 2560 (967) | 3270 (477) | 3330 (541) | 3550 (492) | 3690 (367) | 3070 (323) | 2640 (506) | 2660 (423) |  |
| Maternal age |  |  |  |  |  |  |  |  |  |  |
| Mean (SD) | 24.1 (4.11) |  |  |  |  |  |  | 30.0 (3.99) | 24.0 (5.09) | 25.2 (6.21) |
| Maternal weight |  |  |  |  |  |  |  |  |  |  |
| Mean (SD) |  |  |  |  |  |  |  |  |  |  |
| Maternal education<br>(years) |  |  |  |  |  |  |  |  |  |  |
| Mean (SD) | 5.43 (4.10) |  |  |  |  |  |  | 0.873 (2.51) | 2.48 (4.01) | 4.07 (2.95) |
| Number of rooms |  |  |  |  |  |  |  |  |  |  |
| 4+ | 14 (3.75%) |  |  |  |  |  | 78 (36.3%) |  | 323 (46.3%) |  |
| 1 | 202 (54.2%) |  |  |  |  |  | 44 (20.5%) |  | 49 (7.02%) |  |
| 2 | 106 (28.4%) |  |  |  |  |  | 54 (25.1%) |  | 145 (20.8%) |  |
| 3 | 51 (13.7%) |  |  |  |  |  | 39 (18.1%) |  | 181 (25.9%) |  |
| Number of children<br><5yrs |  |  |  |  |  |  |  |  |  |  |
| 1 |  |  |  |  |  |  |  |  |  |  |
| 2+ |  |  |  |  |  |  |  |  |  |  |
| Improved sanitation |  |  |  |  |  |  |  |  |  |  |
| 1 |  |  |  |  |  |  | 201 (93.5%) |  |  |  |
| 0 |  |  |  |  |  |  | 14 (6.51%) |  |  |  |
| Food security level |  |  |  |  |  |  |  |  |  |  |
| Food Secure |  |  |  |  |  |  |  |  | 479 (71.1%) |  |
| Mildly Food Insecure |  |  |  |  |  |  |  |  | 106 (15.7%) |  |
| Food Insecure |  |  |  |  |  |  |  |  | 89 (13.2%) |  |

|  | IRC, India | JiVitA-3,<br>Bangladesh | JiVitA-4,<br>Bangladesh | Keneba,<br>Gambia | LCNI-5,<br>Malawi | MAL-ED,<br>Bangladesh | MAL-ED,<br>Brazil | MAL-ED,<br>India | MAL-ED,<br>Nepal | MAL-ED,<br>Peru | MAL-ED, South<br>Africa |
| --- | --- | --- | --- | --- | --- | --- | --- | --- | --- | --- | --- |
|  | (N=410) | (N=28300) | (N=5449) | (N=2954) | (N=840) | (N=265) | (N=233) | (N=251) | (N=240) | (N=303) | (N=314) |
| Sex |  |  |  |  |  |  |  |  |  |  |  |
| Female | 185<br>(45.1%) | 13785 (48.7%) | 2728 (50.1%) | 1426 (48.3%) | 421 (50.1%) | 136 (51.3%) | 113 (48.5%) | 138 (55.0%) | 110 (45.8%) | 143 (47.2%) | 159 (50.6%) |
| Male | 225<br>(54.9%) | 14515 (51.3%) | 2721 (49.9%) | 1528 (51.7%) | 419 (49.9%) | 129 (48.7%) | 120 (51.5%) | 113 (45.0%) | 130 (54.2%) | 160 (52.8%) | 155 (49.4%) |
| Birthweight |  |  |  |  |  |  |  |  |  |  |  |
| Mean (SD) | 2890 (448) | 2550 (428) | 2990 (823) | 2970 (421) |  | 2800 (412) | 3340 (483) | 2890 (442) | 2980 (390) | 3130 (430) | 3130 (464) |
| Maternal age |  |  |  |  |  |  |  |  |  |  |  |
| Mean (SD) | 23.7 (3.68) | 21.4 (5.27) |  | 27.4 (7.16) | 26.8 (7.87) | 24.8 (4.99) | 24.8 (5.53) | 23.9 (4.10) | 26.4 (3.76) | 24.2 (6.06) | 26.4 (6.86) |
| Maternal weight |  |  |  |  |  |  |  |  |  |  |  |
| Mean (SD) |  | 43.7 (6.13) |  |  | 50.9 (6.66) | 49.6 (8.54) | 61.9 (11.8) | 50.4 (9.37) | 56.3 (8.27) | 55.5 (8.95) | 67.5 (14.9) |
| Maternal education<br>(years) |  |  |  |  |  |  |  |  |  |  |  |
| Mean (SD) | 5.06 (4.61) | 5.06 (3.82) | 7.51 (4.82) |  | 3.54 (3.34) | 5.63 (2.58) | 9.18 (2.81) | 7.88 (3.17) | 8.77 (3.44) | 7.81 (2.79) | 10.3 (1.94) |
| Number of rooms |  |  |  |  |  |  |  |  |  |  |  |
| 4+ | 17 (4.17%) | 1181 (4.18%) | 286 (5.26%) |  |  | 12 (4.96%) | 127 (60.5%) | 25 (10.6%) | 131 (55.5%) | 139 (51.1%) | 196 (76.3%) |
| 1 | 185<br>(45.3%) | 16754 (59.3%) | 3196 (58.8%) |  |  | 152 (62.8%) | 4 (1.90%) | 84 (35.7%) | 52 (22.0%) | 19 (6.99%) | 14 (5.45%) |
| 2 | 170<br>(41.7%) | 7605 (26.9%) | 1389 (25.6%) |  |  | 50 (20.7%) | 20 (9.52%) | 78 (33.2%) | 31 (13.1%) | 52 (19.1%) | 22 (8.56%) |
| 3 | 36 (8.82%) | 2723 (9.63%) | 562 (10.3%) |  |  | 28 (11.6%) | 59 (28.1%) | 48 (20.4%) | 22 (9.32%) | 62 (22.8%) | 25 (9.73%) |
| Number of children <5yrs |  |  |  |  |  |  |  |  |  |  |  |
| 1 | 89 (21.7%) | 18200 (64.4%) |  |  | 391 (48.2%) |  |  |  |  |  |  |
| 2+ | 321<br>(78.3%) | 10066 (35.6%) |  |  | 421 (51.8%) |  |  |  |  |  |  |
| Improved sanitation |  |  |  |  |  |  |  |  |  |  |  |
| 1 |  | 19609 (69.4%) | 4191 (77.1%) |  | 3 (0.369%) | 204 (84.3%) | 206 (98.1%) | 108 (46.4%) | 235 (99.6%) | 65 (24.7%) | 4 (1.60%) |
| 0 |  | 8655 (30.6%) | 1242 (22.9%) |  | 810 (99.6%) | 38 (15.7%) | 4 (1.90%) | 125 (53.6%) | 1 (0.424%) | 198 (75.3%) | 246 (98.4%) |
| Food security level |  |  |  |  |  |  |  |  |  |  |  |
| Food Secure |  | 8711 (50.0%) | 2751 (50.9%) |  |  | 161 (83.0%) | 3 (2.33%) | 190 (89.6%) | 94 (73.4%) | 27 (23.9%) | 132 (56.7%) |
| Mildly Food Insecure |  | 5793 (33.3%) | 1951 (36.1%) |  |  | 4 (2.06%) | 11 (8.53%) | 5 (2.36%) | 15 (11.7%) | 29 (25.7%) | 19 (8.15%) |
| Food Insecure |  | 2910 (16.7%) | 703 (13.0%) |  |  | 29 (14.9%) | 115 (89.1%) | 17 (8.02%) | 19 (14.8%) | 57 (50.4%) | 82 (35.2%) |

|  | MAL-ED,<br>Tanzania | NIH-Birth,<br>Bangladesh | NIH-Crypto,<br>Bangladesh | PROBIT,<br>Belarus | PROVIDE,<br>Bangladesh | ResPak,<br>Pakistan | SAS-CompFeed,<br>India | SAS-FoodSuppl,<br>India | Tanzania Child 2 | TDC, India | ZVITAMBO,<br>Zimbabwe |
| --- | --- | --- | --- | --- | --- | --- | --- | --- | --- | --- | --- |
|  | (N=262) | (N=629) | (N=758) | (N=17046) | (N=700) | (N=284) | (N=1535) | (N=418) | (N=2400) | (N=160) | (N=14104) |
| Sex |  |  |  |  |  |  |  |  |  |  |  |
| Female | 133 (50.8%) | 297 (47.2%) | 381 (50.3%) | 8217 (48.2%) | 332 (47.4%) | 136 (47.9%) | 702 (45.7%) | 223 (53.3%) | 1184 (49.3%) | 75 (46.9%) | 6847 (48.5%) |
| Male | 129 (49.2%) | 332 (52.8%) | 377 (49.7%) | 8829 (51.8%) | 368 (52.6%) | 148 (52.1%) | 833 (54.3%) | 195 (46.7%) | 1216 (50.7%) | 85 (53.1%) | 7257 (51.5%) |
| Birthweight |  |  |  |  |  |  |  |  |  |  |  |
| Mean (SD) | 3180 (452) | 2710 (414) | 2800 (410) | 3440 (419) | 2780 (371) | 2930 (525) | 2750 (516) |  | 3230 (474) | 2910 (450) | 2970 (456) |
| Maternal age |  |  |  |  |  |  |  |  |  |  |  |
| Mean (SD) | 28.4 (6.67) | 25.0 (5.32) | 24.3 (4.68) | 24.9 (4.92) | 24.7 (4.65) |  | 23.6 (4.08) | 25.0 (4.63) | 26.4 (5.04) |  | 24.5 (5.30) |
| Maternal weight |  |  |  |  |  |  |  |  |  |  |  |
| Mean (SD) | 55.7 (9.11) | 48.0 (8.51) | 52.1 (9.17) | 66.2 (12.5) | 49.3 (9.41) |  | 47.6 (7.05) |  | 62.2 (11.7) |  | 60.0 (11.1) |
| Maternal education (years) |  |  |  |  |  |  |  |  |  |  |  |
| Mean (SD) | 6.17 (1.79) | 3.76 (3.45) | 5.77 (3.63) | 13.0 (1.40) | 4.33 (3.62) |  | 4.42 (4.52) | 1.25 (2.70) | 7.90 (2.27) |  | 9.66 (1.95) |
| Number of rooms |  |  |  |  |  |  |  |  |  |  |  |
| 4+ | 108 (43.2%) |  | 95 (12.5%) |  | 23 (3.29%) |  |  |  |  | 5 (3.13%) |  |
| 1 | 13 (5.20%) |  | 368 (48.5%) |  | 507 (72.4%) |  |  |  |  | 91 (56.9%) |  |
| 2 | 63 (25.2%) |  | 191 (25.2%) |  | 108 (15.4%) |  |  |  |  | 49 (30.6%) |  |
| 3 | 66 (26.4%) |  | 104 (13.7%) |  | 62 (8.86%) |  |  |  |  | 15 (9.38%) |  |
| Number of children <5yrs |  |  |  |  |  |  |  |  |  |  |  |
| 1 |  |  | 541 (71.4%) |  | 512 (73.1%) |  |  |  | 1640 (68.6%) |  |  |
| 2+ |  |  | 217 (28.6%) |  | 188 (26.9%) |  |  |  | 749 (31.4%) |  |  |
| Improved sanitation |  |  |  |  |  |  |  |  |  |  |  |
| 1 |  | 242 (38.5%) | 655 (87.7%) |  | 58 (96.7%) |  |  |  |  |  |  |
| 0 |  | 387 (61.5%) | 92 (12.3%) |  | 2 (3.33%) |  |  |  |  |  |  |
| Food security level |  |  |  |  |  |  |  |  |  |  |  |
| Food Secure |  | 95 (15.1%) | 453 (59.8%) |  |  |  |  |  |  |  |  |
| Mildly Food Insecure |  | 492 (78.2%) | 219 (28.9%) |  |  |  |  |  |  |  |  |
| Food Insecure |  | 42 (6.68%) | 86 (11.3%) |  |  |  |  |  |  |  |  |

**Extended Data Table 3.** Countries and survey years included in the analysis of Demographic and Health Survey data

|  |  |
| --- | --- |
| **Africa** |  |
| Gambia | 2013 |
| Malawi | 2015-2016 |
| Tanzania | 2015-2016 |
| South Africa | 2016 |
| Zimbabwe | 2015 |
| **South Asia** |  |
| Bangladesh | 2014 |
| India | 2015-2016 |
| Nepal | 2016 |
| Pakistan | 2017-2018 |
| **Latin America** |  |
| Brazil | 1996 |
| Guatemala | 2014-2015 |
| Peru | 2012 |

